# Defining the molecular correlate of arteriolar hyalinosis in kidney disease progression by integration of single cell transcriptomic analysis and pathology scoring

**DOI:** 10.1101/2023.06.14.23291150

**Authors:** Rajasree Menon, Edgar A. Otto, Laura Barisoni, Ricardo Melo Ferreira, Christine P. Limonte, Bradley Godfrey, Felix Eichinger, Viji Nair, Abhijit S. Naik, Lalita Subramanian, Vivette D’Agati, Joel M. Henderson, Leal Herlitz, Krzysztof Kiryluk, Dennis G. Moledina, Gilbert W. Moeckel, Paul M. Palevsky, Chirag R. Parikh, Parmjeet Randhawa, Sylvia E. Rosas, Avi Z. Rosenberg, Isaac Stillman, Robert Toto, Jose Torrealba, Miguel A. Vazquez, Sushrut S. Waikar, Charles E. Alpers, Robert G. Nelson, Michael T. Eadon, Matthias Kretzler, Jeffrey B. Hodgin, Kidney Precision Medicine Project (KPMP), Nephrotic Syndrome Study Network (NEPTUNE)

## Abstract

Arteriolar hyalinosis in kidneys is an independent predictor of cardiovascular disease, the main cause of mortality in chronic kidney disease (CKD). The underlying molecular mechanisms of protein accumulation in the subendothelial space are not well understood. Using single cell transcriptomic data and whole slide images from kidney biopsies of patients with CKD and acute kidney injury in the Kidney Precision Medicine Project, the molecular signals associated with arteriolar hyalinosis were evaluated. Co-expression network analysis of the endothelial genes yielded three gene set modules as significantly associated with arteriolar hyalinosis. Pathway analysis of these modules showed enrichment of transforming growth factor beta / bone morphogenetic protein (TGFβ / BMP) and vascular endothelial growth factor (VEGF) signaling pathways in the endothelial cell signatures. Ligand-receptor analysis identified multiple integrins and cell adhesion receptors as over-expressed in arteriolar hyalinosis, suggesting a potential role of integrin-mediated TGFβ signaling. Further analysis of arteriolar hyalinosis associated endothelial module genes identified focal segmental glomerular sclerosis as an enriched term. On validation in gene expression profiles from the Nephrotic Syndrome Study Network cohort, one of the three modules was significantly associated with the composite endpoint (> 40% reduction in estimated glomerular filtration rate (eGFR) or kidney failure) independent of age, sex, race, and baseline eGFR, suggesting poor prognosis with elevated expression of genes in this module. Thus, integration of structural and single cell molecular features yielded biologically relevant gene sets, signaling pathways and ligand-receptor interactions, underlying arteriolar hyalinosis and putative targets for therapeutic intervention.

## INTRODUCTION

Cardiovascular disease (CVD) is the main cause of mortality in patients with CKD^1, 2^. After adjusting for traditional cardiovascular risk factors, impaired kidney function and albuminuria increase the risk of CVD by two- to four-fold.^3-5^. Chronic changes to the structure and function of the kidney vasculature have long been documented in kidney diseases. The vascular structural changes of arteriosclerosis and arteriolar hyalinosis, accumulation of serum proteins in the subendothelial space, are commonly seen in kidney tissue from patients with hypertension, diabetes, focal segmental glomerulosclerosis, and aging.^6-11^ More recent studies have shown that microvascular damage in kidneys is an independent predictor of disease progression in chronic kidney disease (CKD)^12^ and diabetic kidney disease (DKD).^11, 13^ Furthermore, arteriolar hyalinosis is an independent predictor of cardiovascular events [hazard ratio of 1.99] in fully adjusted models in kidney diseases^14, 15^, suggesting that kidney microvasculature damage is likely an indicator of broader organ vascular damage. The presence of arteriolar hyalinosis has also a local effect on other vascular parameters and kidney structures such as arteriolar and glomerular capillary dilatation, and subsequent development and acceleration of focal segmental glomerulosclerosis (FSGS), suggesting that arteriolar hyalinosis is a morphologic correlate of loss of autoregulation.^7, 9^ Therefore, a comprehensive understanding of the underlying molecular mechanisms associated with vascular damage may contribute to a better understanding of CKD and CVD progression and uncover novel therapeutic targets.

A unique opportunity to define vascular injury at both the structural and molecular level is afforded by the Kidney Precision Medicine Project (KPMP). KPMP is a multicenter research network working towards generating molecular atlases of healthy and diseased kidney by using multiple state-of-the-art omics and imaging technologies.^16^ The KPMP prospectively obtains kidney biopsy tissue, ethically and safely, from participants with CKD and acute kidney injury (AKI) primarily for research purposes.

Single-cell RNA sequencing (scRNAseq) data from KPMP has been used to molecularly define distinct glomerular endothelial cell subpopulations in CKD, investigate the clinical relevance of these subgroups and identify a biomarker (α2 macroglobulin) of poor outcome in FSGS patients.^17^ In the current study scRNAseq and a semi quantitative measure of arteriolar hyalinosis in CKD and AKI from whole slide images (WSIs) of kidney biopsies were used to gain insights into cellular processes associated with vascular structural lesions, endothelial crosstalk within the kidney vasculature, and long-term outcomes of kidney function.

## METHODS

### Human data

Participants were recruited after informed consent and race was self-reported. <u>CKD and AKI</u>: Forty-eight (14 AKI and 34 CKD) KPMP kidney biopsy samples were donated by participants (University of Washington IRB#20190213). Among the CKD biopsies, 26 were from patients with DKD and 8 had hypertensive kidney disease (**Supplementary Table S1**). KPMP biopsy tissues were triaged according to protocol by diagnostic and tissue interrogation cores. The diagnostic biopsy core was processed according to standard of care for histology, immunofluorescence and electron microscopy evaluation. Glass slides from the formalin-fixed and paraffin embedded sections, frozen sections, and plastic sections were sent to the University of Michigan and scanned into WSIS at 40X. The tissue interrogation core was preserved using CryoStor® (Stemcell Technologies), and then processed at the University of Michigan to generate single cell RNA sequencing (scRNAseq) data as described below.

### Human Kidney Transplant data

Single cell data was generated at University of Michigan from living donors and surveillance biopsies at 3- and 12-months post-transplantation from the Human Kidney Transplant Transcriptomic Atlas (University of Michigan IRBMED HUM00150968). The dataset contained 40,254 cells derived from 18 living donors, and 6 samples each from 3- and 12-months surveillance biopsies^18^ that enabled comparisons over time post-transplantation.

### NEPTUNE cohort bulk mRNA sequencing data

Longitudinal clinical data were available for the Nephrotic Syndrome Study Network^19^ (NEPTUNE) cohort (**Supplementary Table S2**). The bulk mRNA sequencing data generated from glomerular and tubulointerstitial compartments of kidney biopsies in NEPTUNE were used to test the association of the arteriolar hyalinosis gene set modules with disease progression using multivariable adjusted cox proportional hazard analyses. NEPTUNE (NCT01209000) is a multi-center prospective study of children and adults with proteinuria recruited at the time of first clinically indicated kidney biopsy. Reporting race and ethnicity in NEPTUNE was mandated by the US National Institutes of Health, consistent with the Inclusion of Women, Minorities, and Children policy.

### DKD data from the American Indian cohort

This American Indian cohort^20^ comprised 111 adult participants with type 2 diabetes who provided a kidney biopsy for a prospective, randomized, placebo-controlled, double-blinded intervention trial (NCT00340678). The bulk mRNA sequencing data generated from these kidney biopsies were originally used to examine the effect of Losartan medication on gene module expression.

### Descriptor scoring

WSI of the formalin-fixed and paraffin-embedded sections from the KPMP cohort were scored for severity of arteriolar hyalinosis using a standardized semiquantitative approach: 0 (none), 1+ (mild), 2+ (moderate), or 3+ (severe) ^21^. Arteriolar hyalinosis was defined as accumulation of glassy deposition of homogenous eosinophilic or PAS-positive material in the subendothelial and/or medial space. WSIs were scored by two kidney pathologists. A consensus score was achieved and used in the analyses. Descriptor scores from 45 out of 48 KPMP disease biopsy samples were available and used in this study (**Supplementary Table S3**).

### Single cell sample processing

Single cell dissociation of the samples was performed using Liberase TL at 37°C. Detailed sample processing protocol can be found in <u>dx.doi.org/10.17504/protocols.io.7dthi6n</u>. The single cell suspension was immediately transferred to the University of Michigan Advanced Genomics Core facility for further processing. The 10X Chromium Single-Cell 3’ Reagent Kit was used for single cell RNA sequencing according to protocol. Sample demultiplexing, barcode processing, and gene expression quantifications were performed with the 10X Genomics Cell Ranger v3 pipeline using the GRCh38 (hg38) human reference genome. This scRNAseq data from KPMP was used to identify gene sets / modules associated with arteriolar hyalinosis. The datasets from the three disease cohorts were then used to further explore the expression patterns of these gene sets.

### Single cell data analysis

Ambient mRNA contamination was corrected using SoupX (v1.5.0)^22^. To reduce doublets or multiplets from the analysis, a cutoff of > 500 and < 5000 genes per cell was used. Single-cell data from kidney samples generated using 10X Chromium Single-Cell 3’ Reagent Kit v3 tend to exhibit high mitochondrial read content per cell. Hence, a cutoff of < 50% mitochondrial reads per cell was applied. Seurat R package was used for data processing ^23^. Read counts of each sample were normalized first, then features that were repeatedly variable across datasets for integration were selected. Using these features, anchors were identified which were then used for integrating the datasets. The integration using reciprocal principal component analysis (‘RPCA’) was performed on the single cell data from the 48 KPMP disease and the 18-living donor (control group) biopsies to generate a single combined dataset. The combined dataset was further processed, involving normalization, scaling, dimensionality reduction by Principal Component Analysis (PCA), Uniform Manifold Approximation and Projection (UMAP), and unsupervised clustering (resolution 0.1). Raw counts of endothelial cluster were extracted from the integrated dataset based on the expression of Endomucin (EMCN), a cell type specific marker. Similar integrated analysis as above on the endothelial cluster were performed to obtain sub-clusters of endothelial cells. A resolution of 0.6 was used in the unsupervised clustering.

### Weighted Gene Co-expression Network Analysis (WGCNA)

Co-expressed gene-set modules in arteriolar endothelial cells were identified using WGCNA R package ^24^. The highly sparse single cell read count matrix was not optimal for WGCNA analysis ^25^. Hence, WGCNA was performed on expression matrix of metacells generated from the single cell expression data. Analysis was constrained to the cellular compartments responsible for arteriolar hyalinosis, i.e., arteriolar and descending vasa recta cells from the 14 AKI and 34 CKD KPMP biopsies. Descending vasa recta is considered as arteriolar microvessels that supply blood flow to the renal medulla ^26, 27^, hence, these cells were also included. Genes expressed in less than 50% of cells of the count matrix were removed from the analysis. For each sample, metacells were generated by aggregating read counts from 5 cells selected randomly. since meta cell transcriptome had lower sparsity compared to single cell data transcriptome ^28^, but with similar sensitivity and resolution. and only a small number of cells were aggregated to form the metacells from the arteriolar subset of endothelial cells. The gene modules identified were then correlated to patient level arteriolar hyalinosis scores as illustrated in WGCNA R package.

### Trajectory analysis

Pseudotime trajectory analyses were performed separately on the afferent and efferent arteriolar endothelial cells using Slingshot R package ^29^. The preprocessing steps included filtering out genes expressed in less than 20 cells, normalization, dimensionality reduction using PCA, and clustering using Gaussian mixture modeling (GMM). Pseudo-time trajectory inference was performed using slingshot wrapper function. Genes that changed their expression significantly over the course of the trajectory were identified using the tradeSeq R package ^30^. Interaction network of the trajectory cluster marker genes were performed using STRING ^31^ and projected as a network by Cytoscape ^32^. Additional analysis for wiki pathway enrichment were done using ClusterProfiler R package^33^.

### Ligand-receptor interactions

Potential cell-to-cell interactions were studied using NicheNetR ^34^. NicheNetR is a method that predicts ligand–target links between interacting cell types by combining their expression data with prior knowledge on signaling and gene regulatory networks. The analysis was focused on interactions with arteriolar cells.

### Spatial analysis

To substantiate cell-cell interactions, spatial transcriptomic data (10x Visium) from 6 KPMP participants with CKD and available arteriolar hyalinosis descriptor scores were downloaded from the KPMP data repository (https://atlas.kpmp.org/repository/). Three had descriptor scores of “0”, two had “3” and one was “2”.

Cell type transfer scores for each spot were estimated through the transfer anchors method ^35^ (Seurat V4) using the integrated kidney single cell object as a reference to identify the neighborhood/cellular niches in the spatial data. Cell type proportion for each spot was estimated from their relative transfer scores. Niches were defined by clustering spots based on cell type proportions with usual clustering steps in Seurat pipelines^36^.

STUtility R package ^37^ was used to integrate the 6 spatial datasets to corroborate the ligand-receptor interactions. After integration, clusters were annotated using the top markers. Since, afferent and efferent arteriole cell types did not form separate clusters, spots with the expression of afferent arteriole, efferent arteriole, and glomerular endothelial cell type markers: gap junction protein alpha 4 (GJA4) for afferent arteriole; solute carrier family 6 member 6 (SLC6A6) for efferent arteriole; and sclerostin (SOST) for glomerular endothelial were manually annotated as three additional cell types. Glomerular endothelium was included as a separate cluster to study the potential interaction of this cell type due to its proximity with arterioles. Neighborhood analysis was performed using STUtility R package. Next, for investigating the co-localization of each ligand-receptor pair from the top 29 *bona fide* interactions identified using NichenetR, a subset of the integrated dataset was created that contained the spots from afferent / efferent arterioles and the neighboring spots from interacting cell type in which the ligand of interest was expressed. Using this subset, differential expression of the ligand and receptor were performed between samples with and without arteriolar hyalinosis (“0” vs “2 and 3” descriptor score).

## RESULTS

### Single cell analysis

Raw counts from 10,366 endothelial cells were extracted from the integrated single cell dataset from 48 patient and 18 living donor biopsy samples. Further, sub-clustering of the endothelial cells resulted in 10 endothelial cell types that were annotated using markers from Dumas et al^38^ (**Figure 1a, 1b**). The clusters identified include glomerular endothelial (GC-EC), afferent arteriolar/arterial (EC-AA/LA), efferent arteriolar (EC-EA), ascending vasa recta (EC-AVR), descending vasa recta (EC-DVR), peritubular capillary (EC-PTC), post capillary venules (EC-PCV), lymphatic endothelial (EC-LYM), proliferating endothelial (cycEC) and a cluster with high expression of both endothelial and vascular smooth muscle cell markers (EC-VSMC).

**Figure 1:**
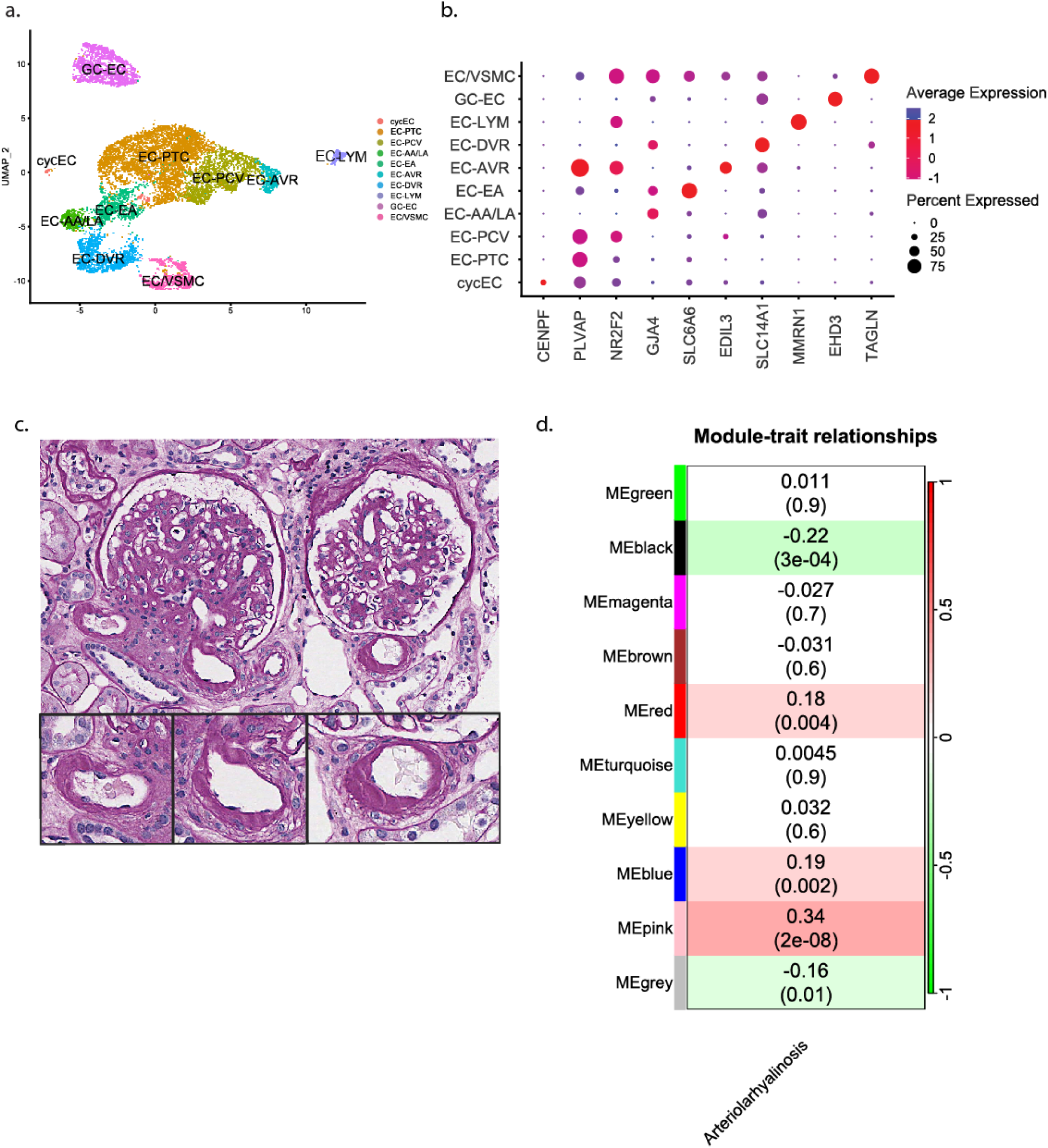
Kidney endothelial clusters and weighted gene correlation network analysis of endothelial gene expression data. **Figure 1a** shows the 10 clusters identified from the sub-clustering of 10,366 endothelial cells from the single cell data of 48 KPMP and 18 living donor kidney biopsies. The clusters identified include glomerular endothelial (GC-EC), afferent arteriolar/arterial (EC-AA/LA), efferent arteriolar (EC-EA), ascending vasa recta (EC-AVR), descending vasa recta (EC-DVR), peritubular capillary (EC-PTC), post capillary venules (EC-PCV), lymphatic endothelial (EC-LYM), proliferating endothelial (cycEC) and a cluster with high expression of both endothelial and vascular smooth muscle cell markers (EC-VSMC). **Figure 1b** provides the dot plot with the cell type specific markers for the 10 sub-clusters identified. **Figure 1c** illustrates a tissue section from the KPMP CKD cohort stained with periodic acid Schiff. Two glomeruli with diabetic kidney disease are represented. The afferent and efferent arterioles of the glomerulus on the left display moderate hyalinosis, the glomerulus on the right display moderate hyalinosis in the arteriolar wall. **Figure 1d** is the results of weighted gene correlation network analysis (WGCNA). The matrix of module-trait relationship reports Kendall’s correlation coefficients, and its corresponding p values, between the eigengene value of each module and the arteriolar hyalinosis. Three modules (blue, pink, and red) show significant correlation with the trait studied, arteriolar hyalinosis.

### Arteriolar hyalinosis scores

Arteriolar hyalinosis was detected to varying extent in afferent and efferent arterioles of the glomeruli in KPMP biopsies (**Figure 1c, Supplementary Table S3**). Examples of moderate hyalinosis in arterioles and arteriolar wall are presented in **Figure 1c**.

### WGCNA analysis

The WGCNA analysis on the metacell expression data of the arteriolar endothelial cells yielded three gene sets/modules (blue, pink, and red) with 28, 11 and 12 genes, respectively, positively associated (p<0.05) with arteriolar hyalinosis (**Figure 1d, Supplementary Table S4**). Positive correlation with arterial hyalinosis persisted for 33 of the 51 genes from these three modules (**Supplementary Table S4**). At the single cell level, the pink module was enriched in the efferent arteriole (EC-EA), red module in the afferent arteriole (EC-AA/LA) and blue module in efferent arteriole, glomerular endothelial, and peritubular capillary cells (**Figure 2a**). Further analyses were focused on EC-EA and EC-AA/LA cells with significant enrichment in all three modules. The density plots of blue, pink, and red module scores in living donor and kidney disease biopsy samples that were down-sampled to a total of 250 arteriolar (afferent and efferent endothelial) cells per group are shown in **Supplementary Figure S1**. The plots show an enrichment of the blue and pink gene modules in kidney disease samples compared to living donor samples.

**Figure 2:**
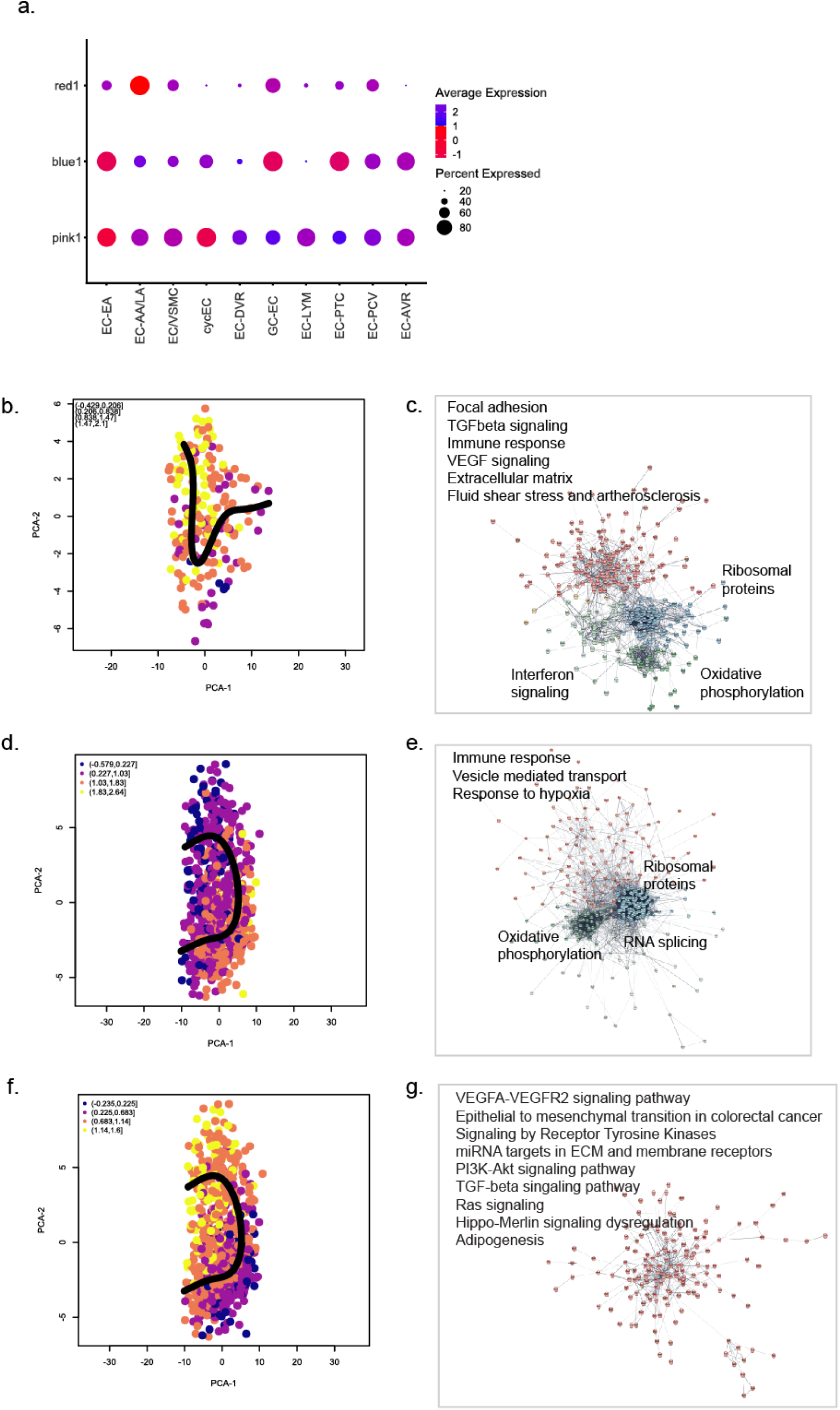
Gene modules associated with arteriolar hyalinosis. **Figure 2a** is the dot plot showing the enrichment of the three gene set modules identified by WGCNA analysis as associated with arteriolar hyalinosis in the 10 endothelial sub clusters. Pink module was most enriched in the EC-EA, blue module was enriched in EC-EA, GC-EC and EC-PTC. The red module was most enriched in the afferent/large artery (EC-AA/LA) endothelial cluster. **Figure 2b** is the pseudo-time trajectory generated for afferent arteriolar cells using Sling shot R package. Enrichment of the red gene set module is observed at the end of the trajectory. **Figure 2c** is the network generated using STRING for the marker genes for the group of cells at the end of the trajectory that were enriched for red module score. The top significant processes for these markers are shown by the network hubs. **Figure 2d** is the pseudo-time trajectory of efferent arteriolar cells showing enrichment of the pink module score. **Figure 2e** shows the STRING network for the marker genes for the cells enriched for pink module genes. **Figure 2f** is the pseudo-time trajectory of efferent arteriolar cells showing enrichment of the blue module genes score. **Figure 2g** shows the top significant processes and the STRING network of the marker genes for the cells with high blue module score at the end of the efferent arteriole trajectory.

### Pseudo-time trajectory analysis

Pseudo-time trajectory analyses showed changes in expression patterns of blue, pink, and red module genes in EC-AA/LA and EC-EA cells in CKD (**Figures 2b-g**). Clustering by GMM produced two distinct clusters along the trajectory of EC-AA/LA. Cells with high red module scores were enriched in the smaller cluster at the end of the EC-AA/LA trajectory (**Figure 2b**). **Supplementary Figure S2a** shows the enriched terms for the genes that were differentially over-expressed at the end compared to the start of the trajectory. The interaction network of the marker genes for the cluster enriched for red module score **(Figure 2c)** consisted of four sub-networks. Two of the sub-networks were hubs with genes enriched for oxidative phosphorylation and ribosomal proteins. The other two sub-networks were enriched for processes including interferon signaling, extracellular matrix, fluid shear stress and atherosclerosis, focal adhesion, immune response and VEGF signaling.

The EC-EA pseudo-time trajectory suggested an initial increase in the expression of pink module genes (**Figure 2d**) followed by an increase in the expression of blue module genes (**Figure 2f**). **Supplementary Figure S2b** shows the enriched terms between the cells with high pink module scores and the cells at the end of the trajectory with high blue module scores. The network of the markers for the cluster with high pink module score included two hubs with genes involved in ribosomal processes and oxidative phosphorylation; in addition, the network included genes enriched in immune response, vesicle mediated transport and focal adhesion (**Figure 2e**). The markers for the cluster with a high blue module score were involved in signaling processes including Hippo-Merlin, RAS, receptor tyrosine kinase, phosphoinositide 3-kinase (PI3K)–AKT, transforming growth factor beta (TGFβ) and vascular endothelial growth factor (VEGF) pathways (**Figure 2g**).

FSGS, NOTCH regulation of endothelial thickening, BMP signaling, adipogenesis, and VEGF signaling were among the significantly enriched wiki pathways for the genes that were differentially expressed along the distinct regions of the trajectory **(Supplementary Tables S5 and S6)**.

### Ligand-receptor interactions

Among the top 50 ligands identified using NicheNetR when arteriolar cells (EC-EA, EC-AA/LA) were set as receiving cells, 29 *bona fide* ligands (literature and publicly available databases) were included (**Figure 3a**). **Figure 3b** shows the expression pattern of these 29 ligands in cell types identified from CKD and AKI samples. Therefore, podocytes, proximal, parietal epithelial, myeloid, T and interstitial cells, were predicted for crosstalk with arteriolar cells. **Supplementary Figure S2c** shows the predicted targets of the 29 ligands among 33 module genes.

**Figure 3:**
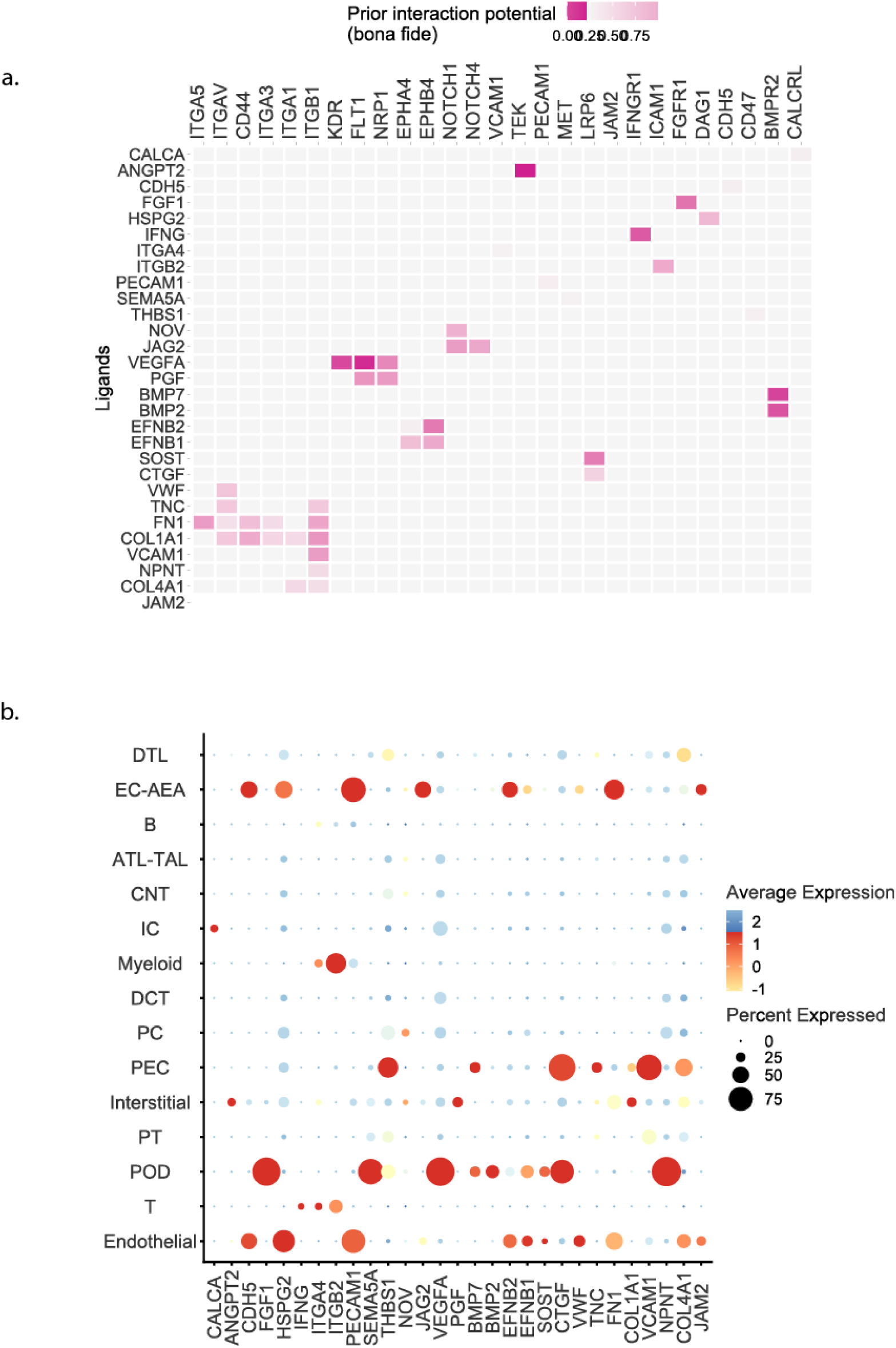
Ligand receptor analysis. **Figure 3a** shows the 29 *bona fide* ligands (literature and publicly available databases) among top 50 identified by NichenetR analysis in which the arteriolar cells were set as receptor cells. The corresponding receptors expressed in the arteriolar cells that potentially interacted with the ligands are shown in the x axis **Figure 3b** is the dot plot showing the expression pattern of the top 29 *bona fide* ligands in the cell types identified in the single cell data from the 48 KPMP kidney biopsies.

Spatial transcriptomic data generated from the KPMP CKD participants support several of the potential ligand-receptor interactions observed in the above NichenetR analysis (**Figure 4).**

**Figure 4:**
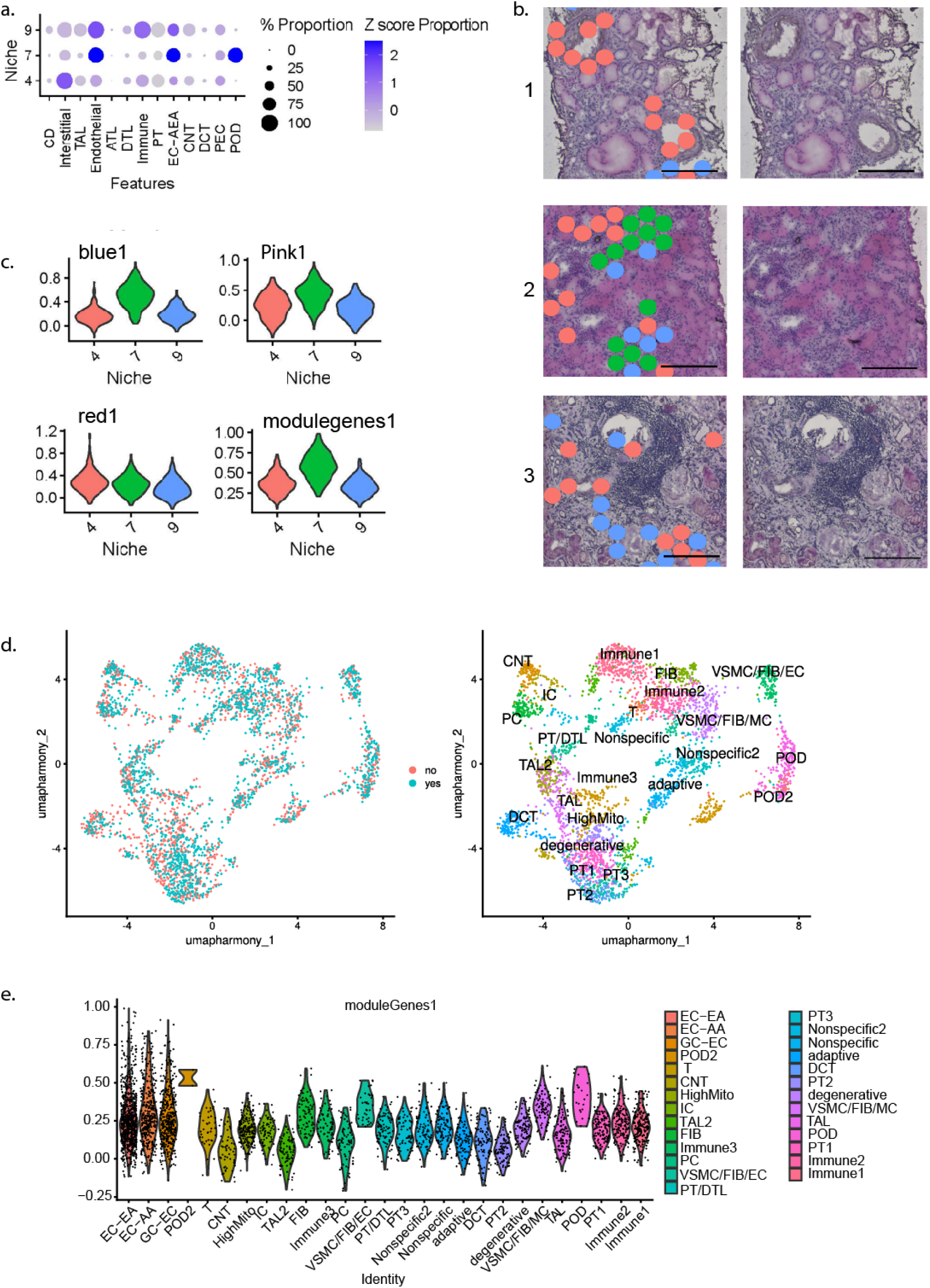
Integrated 10X Visium spatial transcriptomic analysis. **Figure 4a** Dot plot showing the endothelial cell expression in the 3 cell niches identified from the clustering based on the cell type distribution in the spots. Niche 4 represents endothelial cells colocalizing with interstitial cells, 7 has a large proportion of podocytes, indicating glomerular endothelial spots, and 9 is associated with immune cells. **Figure 4b** shows sections from 3 biopsy areas displaying niche 4 localizing around a vessel, niche 7 over glomeruli, and niche 9 is present surrounding a tissue with extensive immune infiltration. **Figure 4c** Violin density plots showing the expression of blue, pink, red and combined module scores (generated from the combined set of genes in the blue, pink and res modules) in the 3 niches with endothelial cells. **Figure 4d** shows the Uniform Manifold Approximation and Projection (UMAP) from the integrated analysis of spatial transcriptomic data from six CKD patients with descriptor scores for arteriolar hyalinosis. Three samples with scores ≥2 was grouped as ‘yes hyalinosis’ and three samples with a score of 0 were termed ‘no hyalinosis’. The first umap shows the integration of all spots from the 6 datasets. The second umap shows the clusters identified from the integration analysis. Clusters were annotated based on top markers. **Figure 4e** Violin plot showing the relative expression pattern of the arteriolar markers (GJA4 and SLC6A6) and the combined module gene score (calculated at spot level) for the clusters identified from the integrated spatial data analysis. The afferent and efferent arteriolar clusters (EC-AA, EC-EA) and the glomerular endothelial cluster (GC-EC) were created by grouping the spots expressing the markers GJA4, SLC6A6 and SOST, respectively.

By clustering the spots based on the cell type distribution, sixteen spatially localized niches were identified (**Supplementary Figure S3**). Three niches were enriched for endothelial cell types (**Figure 4a**). Niche 4 represented endothelial cells co-localized with interstitial cells, 7 had a large proportion of podocytes, indicating glomerular endothelial spots, and 9 was associated with immune cells. Selected biopsy areas display niche 4 localizing around a vessel, niche 7 over glomeruli, and niche 9 is present surrounding a tissue with extensive immune infiltration (**Figure 4b**). The three gene modules blue, pink, and red were highly expressed in those spots (**Figure 4c**).

For the integrated analysis of spatial transcriptomic data from 6 patients with CKD (**Figure 4d),**. of the top 5 genes from all clusters and high expression of EC-AA and EC-EA markers (GJA4 and SLC6A6) are shown in **Supplementary Figure S4**. The endothelial clusters had higher combined module scores (**Figure 4e)**. The ligand-receptor pairs where the expression of either the ligand, the receptor, or both, were higher (nominal p value < 0.05) in samples with arteriolar hyalinosis (descriptor score ≥2 vs 0) were considered to be associated with arteriolar hyalinosis (**Table 1).** The significant co-localized ligand-receptor pairs implied interactions between afferent and efferent arteriolar cells and other cell types including podocytes, interstitial/vascular smooth muscle cells, other endothelial cells, parietal, and immune cells.

**Table 1:** Ligand - receptors interactions. (identified from NichenetR analysis) in which either the ligand or receptor is significantly over-expressed (nominal p value < 0.05) in samples with arteriolar hyalinosis score >0 from the integrated Visium spatial transcriptomic data analysis. The significant p values are in parentheses.

| Ligand from interacting cell type | Corresponding receptor expressed in afferent arterioles |
| --- | --- |
| **Podocyte** |  |
| NPNT (p < 0.008) | ITGB1 (p < 0.039) |
| CCN2 (p < 0.043) | LRP6 |
| **VSMC/FIB/MC** |  |
| COL1A1 (0.004) | CD44 (p < 0.028) |
| **GC-EC** |  |
| FN1 | ITGA5 (p < 0.001) |
| HSPG2 (P < 0.0023) | DAG1 |
| VWF (p < 0.0009) | ITGAV (p < 0.016) |
| **Immune** |  |
| ITGB2 (p < 0.00006) | ICAM1(p < 0.006) |

| Ligand from interacting cell type | Corresponding receptor expressed in efferent arterioles |
| --- | --- |
| **Podocyte** |  |
| FGF1 (p < 0.004) | FGFR1 |
| CCN2 (p < 0.015) | LRP6 |
| **Parietal epithelial** |  |
| COL4A1 (p < 0.06) | ITGA1 |
| VCAM1 (p < 0.004) | ITGB1 |
| **VSMC/FIB/MC** |  |
| PGF | FLT1 (p < 0.028) |
| ANGPT2 | TEK (p < 0.002) |
| **GC-EC** |  |
| EFNB1 (p < 0.00001) | EPHA2 |
| FN1 | ITGB1 (p < 0.028) |
| HSPG2 (p < 0.00001) | DAG1) |
| VWF (p < 0.001) | ITGAV (p < 0.1) |
| JAG2 | NOTCH1 (p < 0.001) |
| **Immune** |  |
| ITGB2 (p < 0.03) | ICAM1(p < 0.0002) |

### Association of co-expressed gene modules with disease progression

To determine whether the blue, pink, and red gene modules contribute to CKD, the association of module scores with disease progression within the Nephrotic Syndrome Study Network (NEPTUNE) ^19^ cohort of 216 adult patients were assessed. All three module scores were significantly associated with disease progression in the unadjusted and adjusted (for age, sex, and race) analyses **(Table 2)**. Pink module scores in both glomerular and tubulointerstitial fractions were associated with disease progression even after adjusting for baseline eGFR (p<0.05) while blue and red module scores were not.

**Table 2:**
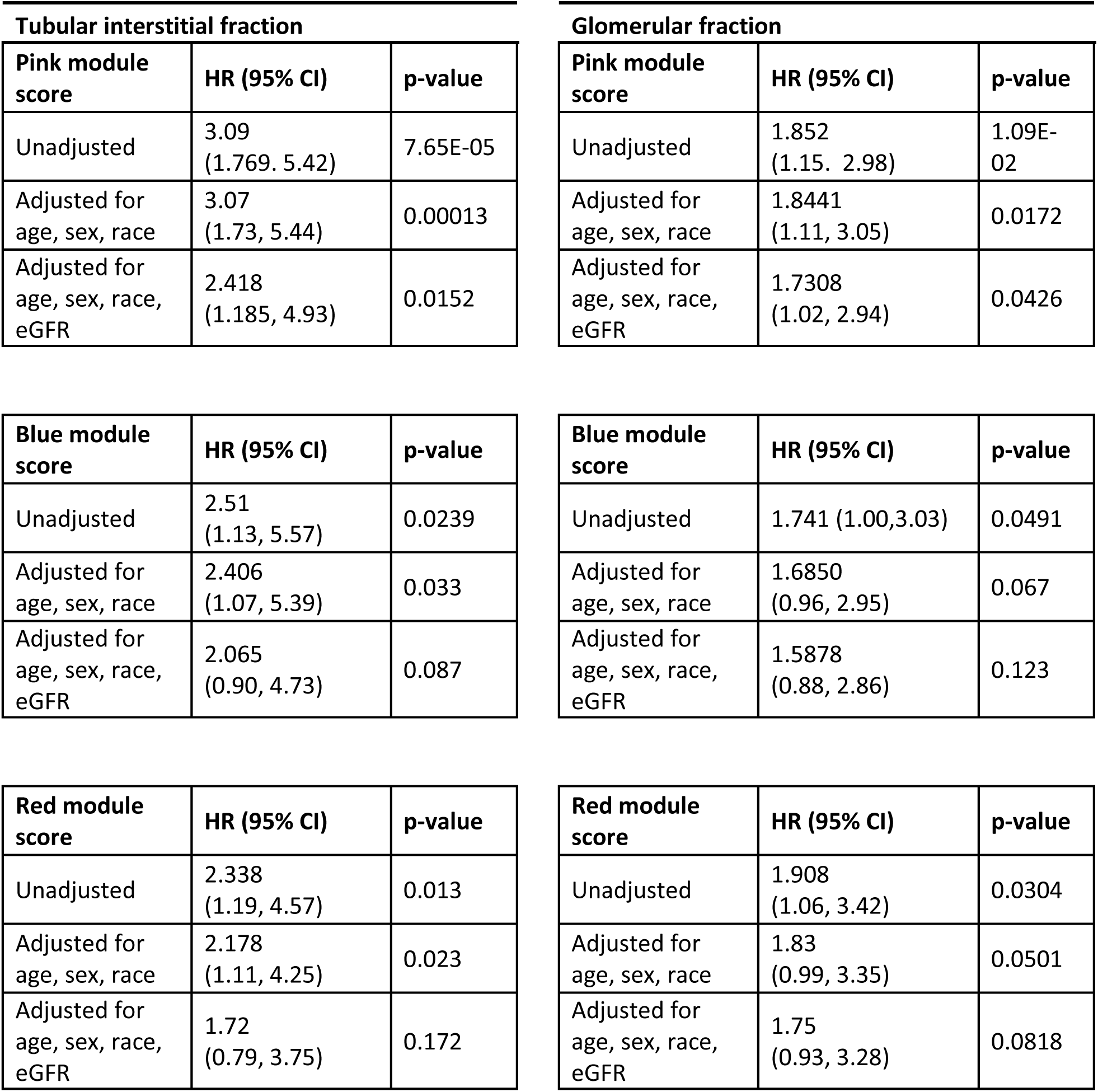
Neptune cohort patients. Adult patients (n=216) with longitudinal data from time of biopsy and 30 events. Adjusted models can only include up to 4 covariates.

### Medication effect on the co-expressed gene modules

Figure 5a shows the effect of RAAS blockade on blue and pink module gene expression in the efferent arteriolar cells in KPMP patients with CKD. As there were not enough endothelial cells to separately analyze the afferent and efferent cell types, we grouped them together as arteriolar cells. To keep the same number of cells in the groups with and without RAAS blockade, we down-sampled to 100 arteriolar cells each. We observe a reduction in pink module gene set expression in patients with RAAS blockade medication compared to those without. The second dataset used to examine the medication effect was the bulk mRNA dataset from 48 participants in the American Indian cohort ^39^. Samples available from 31 patients treated with Losartan showed significant decrease (p<0.05) in pink module gene score compared to the 17 patients from the placebo arm of the trial (Figure 5b).

**Figure 5:**
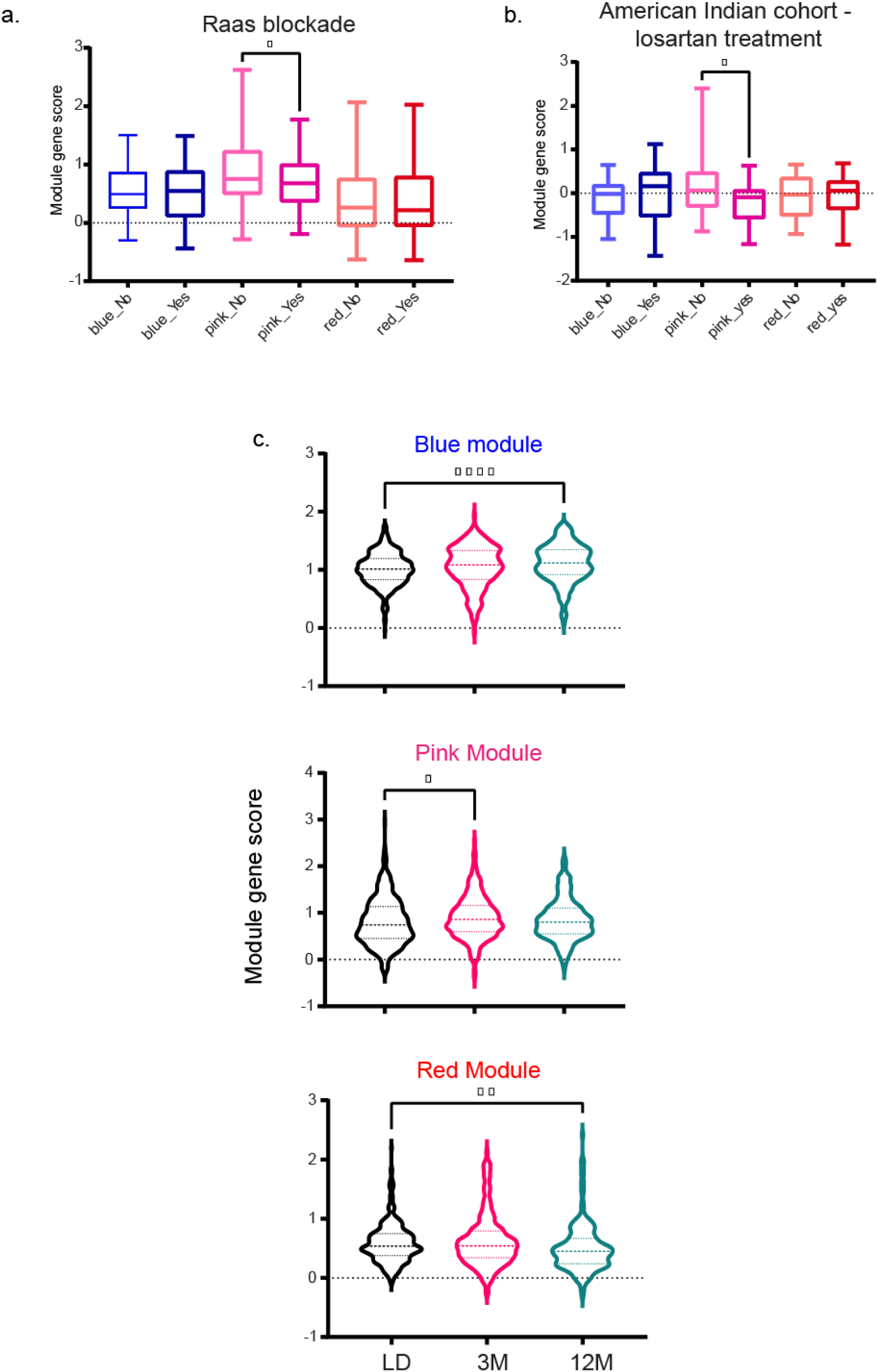
Arteriole enriched gene module expression. **Figure 5a** shows the box plots of blue and pink module scores in the arteriolar cells of KPMP CKD samples with and without RAAS blockade medication. Each sample group was down sampled to 100 cells each. Pink module score showed a significant reduction with RAAS blockade **Figure 5b** shows the box plots of blue and pink module scores in American Indian cohort samples with and without Losartan medication. The plot suggests that Losartan medication resulted in lower pink module gene expression (*p<0.05). **Figure 5c** is the violin plot with blue, pink and red module expression in the arteriolar cells in living donors, and in surveillance biopsies at 3-month and 12-month periods.

### Module expression in transplant single cell data

Sequential protocol biopsies and biopsies for cause in individual patients with transplanted kidneys have previously shown an increase in arteriolar lesions over time without being specific to calcineurin inhibitory toxicity^40^. We examined the expression of pink, blue and red gene modules in arteriolar cells from the single cell data generated from living donors and surveillance biopsies at 3-month and 12-month periods. As there were not enough endothelial cells to separately analyze the afferent and efferent cell types, we grouped them together as arteriolar cells; there were 497 cells in living donors, 232 cells in 3-month and 226 cells in the 12-month groups. The blue and red module expression was the lowest in the living donors and highest in the 12-month group (Figure 5c). The pink module was highest in 3-month and decreased in the 12-month group (Figure 5c).

## DISCUSSION

Arteriolar hyalinosis in kidneys is a common vascular lesion seen in diseases involving the renal vasculature, including aging, hypertension, diabetes mellitus, and focal segmental glomerulosclerosis (FSGS)^13^. It is often associated with systemic vascular stress and CVD^14, 15^; however, not much is known about the specific molecules involved in this process. The availability of histopathology and molecular data from different cohorts, enabled investigation of the relationship between vascular structural changes and mechanisms of damage (Figure 6).

**Figure 6:**
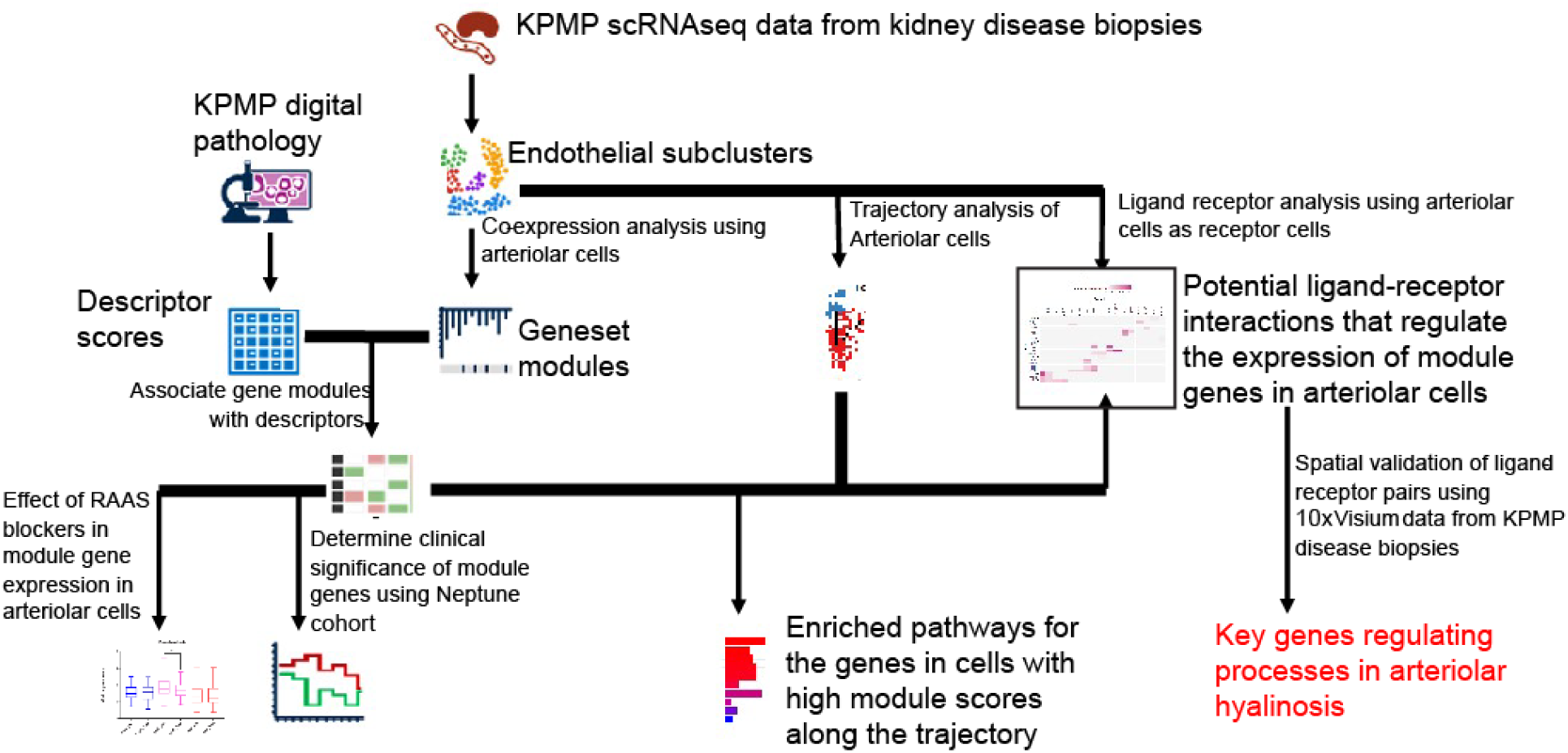
Schema of the analysis processes. The flow diagram describes the various analyses performed in this study integrating pathological descriptor scores and single cell transcriptomic data

By sub-clustering an ample number of endothelial cells (∼10,000 cells) from the single cell data of disease (KPMP) and reference samples (living donor), distinct arteriolar cell clusters could be identified. Three sets of co-expressed gene modules (i.e., blue, pink and red) were enriched in arteriolar cells associated with the corresponding histopathology descriptor scores for arteriolar hyalinosis. Pseudo-time trajectory in arteriolar cells followed by pathway enrichment analysis highlighted signaling pathways known to be implicated in vascular pathogenesis. VEGF signaling was enriched in arteriolar cells with high red and blue gene module scores and NOTCH1 regulation of endothelial cell calcification was enriched for afferent cells with high red module scores. This is consistent with the modulation of NOTCH1 signaling in podocytes alleviating VEGF-mediated diabetic nephropathy in animal models,^41^ and the reported interplay between NOTCH1 and VEGF signaling pathways in vascular disease ^42^.

The enriched processes of the three module gene sets contained immune terms linked to angiogenesis, burn wound healing, interferon 1 signaling, immune response and epithelial to mesenchymal transition in colorectal cancer. The spatial transcriptomic analysis further supported the receptor ligand interaction between immune cells and arteriolar cells (ITGB2 (ligand from immune cells) – ICAM1 (receptor on arteriolar cells). The role of immune related processes in vascular remodeling is under active investigation^43^.

Ligand-receptor analysis suggested cellular crosstalk between parietal epithelial, podocyte, immune and interstitial cells and the vascular beds of the arteriolar cells. Ligands such as VCAM1 and COL1A1 have been previously implicated in kidney disease^44, 45^. The spatial integration analysis further refined the subset of ligand-receptor pairs in the samples with moderate to severe arteriolar hyalinosis.

Through spatial transcriptomic analyses of the CKD samples, increased expression of *CD44*, *ITGA5* and *ITGB1*, receptors of matrix proteins such as collagens (COL1A1, COL4A1) and fibronectins (FN1) were detected in the arteriolar cells of samples with high arteriolar hyalinosis descriptor scores, consistent with known deposition of hyaline into the vascular wall coupled with matrix protein synthesis in arteriolar hyalinosis^46^. Integrins and TGFβ 1 pathway interactions have been described in both angiogenic and pathogenic mechanisms.^47,48^ Interestingly, TGFβ /BMP signaling was enriched in arteriolar cells with high red and blue module gene scores. Several studies have implicated TGFβ as the primary initiator of arteriolar hyalinosis ^49-52^. The expression of bone morphogenetic proteins (BMPs), a family of TGFβ proteins, has been shown to be increased at vascular calcification sites ^53^. Effect of vascular homeostasis disturbance on BMP expression has implicated them in abnormal vascular responses^54^. This study supports the involvement of integrins and TGFβ /BMP pathways in arteriolar hyalinosis processes.

In arteriolar hyalinosis, the muscle layer of the arterioles is thinner ^13^ in the areas of hyaline deposition, likely impairing the ability of the vessel to constrict. Prostaglandin (PG) have a well-established role in maintaining glomerular filtration rate by dilating the afferent arteriole and stimulating renin release, increasing levels of angiotensin II that preferentially constricts the efferent arteriole ^55^. In line with this role, PG metabolism was one of the enriched terms for the red module gene set.

High expression of the ligand CCN2 (CTGF) in podocytes interacted with LRP6 expressed in both afferent and efferent arterioles in the arteriolar hyalinosis samples. Connective growth factors including CCN2 (CTGF) have been implicated in DKD^56^ This study, for the first time, provides insight into their function in the context of arteriolar hyalinosis.

Meanwhile, in the endothelial trajectories, the pink module gene set was enriched “earlier”, i.e., associated with less severe damage, in the trajectory path of the efferent arteriolar cells, whereas the blue module gene set was enriched at the end of the trajectory. Immune response and oxidative phosphorylation pathways, enriched in pink module genes, suggesting a role at an earlier stage in the hyalinosis process. The pink and blue gene set expression pattern observed in the arteriolar cells of the living donors and surveillance transplant biopsies supported this postulation, with pink module genes more elevated in 3-month samples compared to 12-month post-transplantation. The blue module was most enriched in the 12-month group matching its position at the end of the pseudo-time trajectory in CKD.

FSGS was one of the enriched terms linking arteriolar hyalinosis to glomerular sclerosis. This linkage was confirmed by the association of the pink and blue module scores with long-term outcomes of FSGS disease progression in NEPTUNE. The association persisted for the pink module even when adjusting for age, sex, race and baseline eGFR.

RAAS blockade was associated with decreased pink module gene expression in KPMP. This finding was further supported by significant lower pink module activity levels in the exit biopsies of patients in the treatment arm of the randomized, placebo-controlled clinical trial comparing Losartan to placebo in incipient DKD. Enriched oxidative phosphorylation in the cells with high pink module score suggest increased production of free radicals leading to oxidative stress in the arteriolar wall and that this mechanism is effectively targeted by RAAS inhibition. Individuals with CKD are often on several medications given the prevalence of diabetes mellitus, hypertension, and other CVD risk factors. Nevertheless, by combining observational and controlled trial data in the American Indian cohort, effects of an individual drug exposure on gene expression levels were determined.

This study had some limitations. The ligand-receptor analyses only focused on ligands from cells within the kidney. However, biologically relevant ligands are likely to be found in the plasma or circulating immune cells, as well. This will require integration of multi-omics data from blood or plasma, in the future. Another caveat pertains to the cellular heterogeneity captured by spatial transcriptomics. With the current technology, a single spot in spatial data can contain 1 to 30 cells that are from the same or adjacent cell types in the kidney tissue. Thus, interpretation of ligand-receptor interactions can be challenging. However, cell marker filters were used to enrich the spots with arteriolar cells or the interacting cell type. Future studies with spatial transcriptomics at single cell resolution will be more conclusive.

In conclusion, by integrating kidney biopsy single cell and spatial transcriptomics data with pathology descriptor scores, afforded by the KPMP and sister networks, cellular crosstalk in endothelial cells in arteriolar hyalinosis were identified. Novel and known signaling pathways and ligand receptor interactions were not only linked to poor long-term patient outcomes, but also responded to Losartan treatment. This provides the molecular underpinnings of known treatments for vascular disease in CKD in humans while offering novel targets for drug development. This methodological approach opens new frontiers for studying human diseases at the single cell level and paving the way for future mechanistic studies into vascular damage in CKD.

## Data Availability

The KPMP single cell data used in this study is available from https://atlas.kpmp.org/repository/. Other datasets used include transcriptomic data from NEPTUNE, an American Indian cohort and the Human Kidney Transplant Transcriptomic Atlas. Neptune mRNA sequencing and clinical data are controlled access data and will be available to researchers upon request to. Owing to ethical considerations and privacy protection concerns, and to avoid identifying individual study participants in Pima Indian population, the Institutional Review Board of the National Institute of Diabetes and Digestive and Kidney Diseases has stipulated that individual-level gene expression and genotype data from this study cannot be made publicly available for the American Indian cohort. The single cell count matrices for the Human Kidney Transplant Transcriptomic Atlas are available from the Gene Expression Omnibus under accession number GSE 169285.

https://atlas.kpmp.org/repository/

## ACKNOWLEDGEMENTS

We are deeply indebted to the generosity of patients volunteering to donate tissue primarily for research purposes despite no direct immediate benefit to their clinical care. We thank the KPMP clinical coordinators at the recruitment sites for their efforts in patient enrolments and biopsy tissue procurement, pathologists, and the Central Hub for coordinating data collection, storage and making it accessible to the public through the consortium website.

The KPMP data presented here is funded by the following grants from the NIDDK: U2C DK114886, UH3DK114861, UH3DK114866, 2U01DK114866-07, UH3DK114870, UH3DK114908, UH3DK114915, UH3DK114926, UH3DK114907, UH3DK114923 and UH3DK114933. This work was supported in part by the Intramural Research Program at the National Institute of Diabetes and Digestive and Kidney Diseases (DK069062) to RGN, the American Diabetes Association (Clinical Science Award 1-08-CR-42) to RGN, and (DK083912, DK082841, DK020572, DK092926) to RGN, by the extramural research program of the National Institute of Diabetes and Digestive and Kidney Diseases R24 DK082841 ‘Integrated Systems Biology Approach to Diabetic Microvascular Complications’ and P30 DK081943 ‘University of Michigan O’Brien Kidney Translational Core Center’ to MK. The Nephrotic Syndrome Study Network Consortium (NEPTUNE), U54-DK-083912, is a part of the National Institutes of Health (NIH) Rare Disease Clinical Research Network (RDCRN), supported through collaboration between the Office of Rare Diseases Research, National Center for Advancing Translational Sciences and the National Institute of Diabetes and Digestive and Kidney Diseases. Additional funding and/or programmatic support for this project has also been provided by the University of Michigan, the NephCure Kidney International and the Halpin Foundation. A.S.N. receives salary and research support from NIDDK (K23 DK125529). Christine Limonte was supported by the American Kidney Fund’s Clinical Scientist in Nephrology Award. Dr. Vazquez was supported by grants UH3DK114870-03 and UO1DK133091-01 from the National Institute of Diabetes and Digestive and Kidney Diseases.

## List of Supplementary Materials

1. Supplementary acknowledgements
  a. KPMP Contributors List
  b. Members of the Nephrotic Syndrome Study Network (NEPTUNE)
2. Supplementary Figures
  a. Supplementary Figure S1. Expression levels of module scores in disease and living donor biopsies
  b. Supplementary Figure S2. Enriched wiki pathways and regulatory potential of predicted target genes
  c. Supplementary Figure S3. UMAP of the sixteen spatially localized niches in the 10x Visium spots based on clustering by cell type distribution.
  d. Supplementary Figure S4. Integrated unbiased clustering analysis of six CKD visium transcriptomic datasets.
3. Supplementary Tables
  a. Supplementary Table S1. Time of biopsy characteristics of participants in KPMP cohort included in this study.
  b. Supplementary Table S2. Time of biopsy characteristics of participants in Neptune included in this study.
  c. Supplementary Table S3. Distribution of arteriolar hyalinosis descriptor scores.
  d. Supplementary Table S4. Correlation of module genes with arteriolar hyalinosis score.
  e. Supplementary Table S5. Enriched pathways along the pseudotime in afferent arteriolar cells.
  f. Supplementary Table S6. Pathway differences in efferent cells with high blue and pink module scores.

