## Supplementary materials for "Defining the molecular correlate of arteriolar hyalinosis in kidney disease progression by integration of single cell transcriptomic analysis and pathology scoring"

List of Supplementary Materials

1. **Supplementary acknowledgements**
    a. KPMP Contributors List
    b. Members of the Nephrotic Syndrome Study Network (NEPTUNE)
2. **Supplementary Figures**
    **a. Supplementary Figure S1.** Expression levels of module scores in disease and living donor biopsies
    **b. Supplementary Figure S2.** Enriched wiki pathways **and** regulatory potential of predicted target genes
    **c. Supplementary Figure S3.** UMAP of the sixteen spatially localized niches in the 10x Visium spots based on clustering by cell type distribution.
    **d. Supplementary Figure S4.** Integrated unbiased clustering analysis of six CKD visium transcriptomic datasets.
3. **Supplementary Tables**
    a. Supplementary Table S1. Time of biopsy characteristics of participants in KPMP cohort included in this study.
    b. Supplementary Table 2. Time of biopsy characteristics of participants in NEPTUNE included in this study.
    c. Supplementary Table S3. Distribution of arteriolar hyalinosis descriptor scores.
    d. Supplementary Table S4. Correlation of module genes with arteriolar hyalinosis score.
    e. Supplementary Table S5. Enriched pathways along the pseudotime in afferent arteriolar cells.
    f. Supplementary Table S6. Pathway differences in efferent cells with high blue and pink module scores.

**Supplementary acknowledgement**

**KPMP Contributors List**

Altos Labs: Blue Lake, Kun Zhang

Beth Israel Deaconess Medical Center: Stewart Lecker; Alexander Morales; Isaac Stillman

Boston Cell Standards: Steve Bogen

Boston Medical Center: Afolarin A. Amodu; Laurence Beck; Joel Henderson; Titlayo Ilori; Shana Maikhor; Ingrid Onul, Insa Schmidt; Ashish Verma; Sushrut Waikar; Pranav Yadati; Guanghao Yu

Brigham and Women's Hospital: Mia R. Colona; Gearoid McMahon; Astrid Weins

Broad Institute: Nir Hacohen; Anna Greka; Paul J. Hoover; Jamie L. Marshall

Case Western Reserve University: Mark Aulisio; William Bush; Yijiang Chen; Dana Crawford; Anant Madabhushi; Vidya S. Viswanathan

Cleveland Clinic: Lakeshia Bush; Leslie Cooperman; Crystal Gadegbeku; Leal Herlitz; Stacey Jolly;Jane Nguyen; Charles O'Malley; John O'Toole; Ellen Palmer; Emilio Poggio; Kassandra Spates-Harden; John Sedor; Dianna Sendrey; Jonathan Taliercio

Columbia University: Paul Appelbaum; Olivia Balderes; Jonathan Barasch; Cecilia Berroue; Andrew Bomback; Pietro A. Canetta; Vivette D'Agati; Krzysztof Kiryluk; Satoru Kudose; Karla Mehl; Maya Sabatello; Ning Shang

Delft University of Technology: Joana de Pinho Gonçalves; Roy Lardenoije; Lukasz Migas; Raf Van de Plas

Duke University: Laura Barisoni

Harvard University: Helmut Rennke

Icahn School of Medicine at Mount Sinai: Evren Azeloglu; Kirk Campbell; Steven Coca; Cijang He; John He; Srinivas Ravi Iyengar; Seanee Lefferts; Girish Nadkarni; Marissa Patel; Joji Tokita; Stephen Ward; Yuguang Xiong

Indiana University: Abraham Verdoes; Angela Sabo; Daria Barwinska; Debora Lidia Gisch; James Williams; Katherine Kelly; Kenneth Dunn; Mahla Asghari; Michael Eadon; Michael Ferkowicz; Pierre Dagher; Ricardo Melo Ferreira; Seth Winfree; Sharon Bledsoe; Stephanie Wofford; Tarek El-Achkar; Timothy Sutton; William Bowen; Ying-Hua Cheng; Austen Slade; Elizabeth Record; Yinghua Cheng

Indian University at Bloomington: Katy Borner, Bruce Herr; Yashvardhan Jain; Ellen Quardokus

Johns Hopkins University: Mohamed Atta; Lauren Bernard; Steven Menez; Chirag Parikh; Celia Pamela Corona Villalobos; Ashley Wang; Yumeng Wen; Alan Xu

Joslin Diabetes Center: Sarah Chen; Isabel Donohoe; Camille Johansen; Sylvia Rosas; Jennifer Sun

KPMP Patient Partner: Joseph Ardayfio; Jack Bebiak; Taneisha Campbell; Monica Fox; Richard Knight; Robert Koewler; Roy Pinkeney;  John Saul; Anna Shpigel

Northwestern University: Pottumarthi Prasad

Ohio State University: Sethu M. Madhavan; Samir Parikh; Brad Rovin; John P. Shapiro

Pacific Northwest National Laboratory: Christopher Anderton, Jessica Lukowski; Ljiljana Pasa-Tolic; Dusan Velickovic

Parkland Health: George Oliver

Princeton University: Weiguang Mao; Rachel Sealfon; Olga Troyanskaya; Aaron Wong

Seattle Children's Hospital: Ari Pollack

Stanford University: Yury Goltsev

State University of New York at Buffalo: Brandon Ginley; Brendon Lutnick

University of California at San Diego: Kun Zhang

University of California at San Francisco: Kavya Anjani; Zoltan G. Laszik; Tariq Mukatash; Garry Nolan

University of Arizona: David Beyda; Erika Bracamonte; Frank Brosius; Baltazar Campos; Nicole Marquez; Katherine Mendoza; Raymond Scott; Bijin Thajudeen; Rebecca Tsosie; Gregory Woodhead

University of Chicago: Milda Saunders

University of Cincinnati: Rita R. Alloway; Paul J. Lee; Adele Rike; Tiffany Shi; E. Steve Woodle

University of Colorado: Petter Bjornstad, Elena Hsieh; Jessica Kendrick; Laura Pyle; Joshua Thurman; Carissa Vinovskis; Julia Wrobel

University of Florida: Nicholas Lucarelli; Pinaki Sarder

University of Illinois, Chicago: James Bui; Eunice Carmona-Powell; Ron Gaba; Tanika Kelly; James Lash; Natalie Meza; Devona Redmond; Amada Renteria; Ana Ricardo; Suman Setty; Anand Srivastava

University of Michigan: Fadhl Alakwaa; Heather Ascani; Ul Balis; Markus Bitzer; Victoria Blanc; Nikki Bonevich; Ninive Conser; Dawit Demeke; Rachel Dull; Sean Eddy; Renee Frey; John Hartman; Yongqun Oliver He; Jeffrey Hodgin; Matthias Kretzler; Chrysta Lienczewski; Jinghui Luo; Laura Mariani; Phillip McCown; Rajasree Menon;  Viji Nair; Edgar Otto; Rebecca Reamy; Michael Rose; Jennifer Schaub; Becky Steck; Zachary Wright

University of Minnesota: Alyson Coleman; Dorisann Henderson-Brown; Jerica Berge; Maria Luiza Caramori; Oyedele Adeyi; Patrick Nachman; Sami Safadi; Siobhan Flanagan; Sisi Ma; Susan Klett; Susan Wolf; Tasma Harindhanavudhi; Via Rao

University of North Carolina: Peter Bream; Anne Froment; Sara Kelley; Amy Mottl; Prabir Roy-Chaudhury; Evan Zeitler

University of Pittsburgh: Filitsa Bender; Michele Elder; Matthew Gilliam; Daniel E. Hall; John A. Kellum; Raghavan Murugan; Paul Palevsky; Matthew Rosengart; Roderick Tan; Mitchell Tublin; James Winters

University of Texas Health Science Center at San Antonio: Shweta Bansal; Richard Montellano; Annapurna Pamreddy; Kumar Sharma; Manjeri Venkatachalam; Hongping Ye; Guanshi Zhang

University of Texas Southwestern: Mujeeb Basit; Qi Cai; Allen Hendricks; Susan Hedayati; Asra Kermani; Simon C. Lee; Shihong Ma; Richard Tyler Miller; Orson W. Moe; Harold Park; Jiten Patel; Anil Pillai;; Kamalanathan Sambandam; Jose Torrealba; Robert D. Toto; Miguel Vazquez; Nancy Wang; Natasha Wen; Dianbo Zhang

University of Washington: Charles Alpers; Ashley Berglund; Brooke Berry; Kristina Blank; Keith Brown; Jonas Carson; Stephen Daniel; Ian H. de Boer; Ashveena L Dighe; Frederick Dowd; Stephanie M. Grewenow; Jonathan Himmelfarb; Andrew Hoofnagle; Nichole Jefferson; Brandon Larson; Christine Limonte; Robyn McClelland; Sean Mooney; Yunbi Nam; Christopher Park; Jimmy Phuong; Kasra Rezaei; Glenda Roberts; Natalya Sarkisova; Stuart Shankland; Jaime Snyder; Christy Stutzke; Katherine Tuttle; Artit Wangperawong; Adam Wilcox; Kayleen Williams; Bessie Young

Vanderbilt University: Jamie Allen; Richard M. Caprioli; Mark de Caestecker; Katerina Djambazova; Martin Dufresne; Melissa Farrow; Agnes Fogo; Kavya Sharman; Jeffrey Spraggins

Washington University St. Louis: Jeannine Basta; Kristine Conlon; Sabine M. Diettman; Joseph Gaut; Madhurima Kaushal; Sanjay Jain; Amanda Knoten; Brittany Minor; Gerald Nwanne; Anitha Vijayan; Bo Zhang

Yale University: Tanima Arora; Lloyd Cantley; Angela M. Victoria Castro; Vijayakumar Kakade; Gilbert Moeckel; Dennis Moledina; Melissa Shaw; Francis P Wilson

### Members of the Nephrotic Syndrome Study Network (NEPTUNE)

**NEPTUNE Enrolling Centers**
*Atrium Health Levine Children's Hospital, Charlotte, SC*: Susan Massengill*, Layla Lo#
*Cleveland Clinic, Cleveland, OH*: Katherine Dell*, John Sedor**, Stephanie Larson#
*Children's Hospital, Los Angeles, CA*: Ian Macumber* Kevin Lemley*, Silpa Sharma#
*Children's Mercy Hospital, Kansas City, MO*: Tarak Srivastava*, Kelsey Markus#
*Cohen Children's Hospital, New Hyde Park, NY*: Christine Sethna*, Suzanne Vento #
*Columbia University, New York, NY:* Pietro Canetta*, Anup Pradhan#
*Duke University Medical Center, Durham, NC:* Opeyemi Olabasi**, Rasheed Gbadegesin*, Maurice Smith#
*Emory University, Atlanta, GA:* Laurence Greenbaum*, Chia-shi Wang*, Emily Yun#
*The Lundquist Institute, Torrence, CA:* Sharon Adler*, Janine LaPage#
*John H Stroger Cook County Hospital, Chicago, IL:* Amatur Amarah*, Matthew Itteera#
*Johns Hopkins Medicine, Baltimore, MD:* Meredith Atkinson*, Miahje Williams#
*Mayo Clinic, Rochester, MN:* John Lieske*, Marie Hogan**
*Medical University of South Carolina,* David Selewski*, Cheryl Alston#
*Montefiore Medical Center, Bronx, NY:* Frederick Kaskel*, Kim Reidy**, Michael Ross*, Patricia Flynn#
*NIDDK Intramural, Bethesda MD:* Jeffrey Kopp*
*New York University Medical Center, New York, NY:* Laura Malaga-Dieguez*, Olga Zhdanova**, Laura Jane Pehrson#, Melanie Miranda#
*The Ohio State University College of Medicine, Columbus, OH*: Salem Almaani*, Laci Roberts#
*Stanford University, Stanford, CA:* Richard Lafayette*, Shiktij Dave#
*Temple University, Philadelphia, PA:* Iris Lee*, Zoey Pfeffer#
*Texas Children's Hospital at Baylor College of Medicine, Houston, TX*: Shweta Shah*, Aisha Deslandes# #
*University Health Network Toronto:* Heather Reich*, Michelle Hladunewich**, Paul Ling#, Martin Romano#
*University of California at San Francisco, San Francisco, CA*: Paul Brakeman*
*University of Colorado Anschutz Medical Campus, Aurora, CO*: Amber Podoll* Nathan Rogers#
*University of Kansas Medical Center, Kansas City, KS*: Ellen McCarthy*, Elizabeth Landry#
*University of Miami, Miami, FL:* Alessia Fornoni*, Carlos Bidot#
*University of Michigan, Ann Arbor, MI:* M Kretzler*, Laura Mariani*, Zubin Modi*, A Williams#, Meghan Stelzer#
*University of Minnesota, Minneapolis, MN:* Patrick Nachman*, Michelle Rheault*, Jenna Hanson#
*University of North Carolina, Chapel Hill, NC:* Vimal Derebail*, Keisha Gibson*, Anne Froment#
*University of Pennsylvania, Philadelphia, PA:* Lawrence Holzman*, Kevin Meyers**, Krishna Kallem#, Ann Swenson#
*University of Texas San Antonio, San Antonio, TX*: Samin Sharma*
*University of Texas Southwestern, Dallas, TX:* Elizabeth Roehm*, Kamalanathan Sambandam*, Jamie Hellewege
*University of Washington, Seattle, WA:* Ashley Jefferson*, Sangeeta Hingorani**, Katherine Tuttle**§, Linda Manahan #, Emily Pao#, Kelli Kuykendall§
*Wake Forest University Baptist Health, Winston-Salem, NC:* Jen Jar Lin*
*Washington University in St. Louis, St. Louis, MO*: Ellen Cody*

**Data Analysis and Coordinating Center:** Matthias Kretzler*, Laura Barisoni**, Crystal Gadegbeku**, Brenda Gillespie**, Lawrence Holzman**, Laura Mariani**, Zubin Modi**, Matthew G Sampson**, Eloise Salmon**, John Sedor**, Abigail Smith**, Howard Trachtman**, Jarcy Zee**, Gabrielle Alter, Hailey Desmond, Sean Eddy, Damian Fermin, Wenjun Ju, Maria Larkina, Shengqian Li, Shannon Li, Chrysta Lienczewski, Tina Mainieri, Rebecca Scherr, Jonathan Troost, Amanda Williams

**Digital Pathology Committee:** Carmen Avila-Casado (University Health Network, Toronto), Serena Bagnasco (Johns Hopkins University), Clarissa Cassol (Arakana), Lihong Bu (Mayo Clinic), Shelley Caltharp (Emory University), Dawit Demeke (University of Michigan), Brenda Gillespie (University of Michigan), Jared Hassler (Temple University), Leal Herlitz (Cleveland Clinic), Stephen Hewitt (National Cancer Institute), Jeff Hodgin (University of Michigan), Danni Holanda (Arkana), Neeraja Kambham (Stanford University), Kevin Lemley (Children's Hospital of Los Angeles), Laura Mariani (University of Michigan), Nidia Messias (Washington University), Alexei Mikhailov (Wake Forest), Behzad Najafian (University of Washington), Matthew Palmer

*Principal Investigator; **Co-investigator; #Study Coordinator; §Providence Medical Research Center, Spokane, WA

*Last Update: 18 May 2023*

(University of Pennsylvania), Avi Rosenberg (Johns Hopkins University), Virginie Royal (University of Montreal), Barry Stokes (Columbia University), David Thomas (Duke University), Michifumi Yamashita (Cedar Sinai), Hong Yin (Emory University) Jarcy Zee (University of Pennsylvania), Yiqin Zuo (University of Miami) Co-Chairs: Laura Barisoni (Duke University) and Cynthia Nast (Cedar Sinai).

**Supplementary Figure S1. Expression levels of module scores in disease and living donor biopsies.** Density violin plot showing the relative expression levels of the module scores calculated for the blue, pink and red gene-sets in disease and living donor biopsies. The samples were down sampled to a total of 250 arteriolar (afferent ad efferent endothelial) cells per group

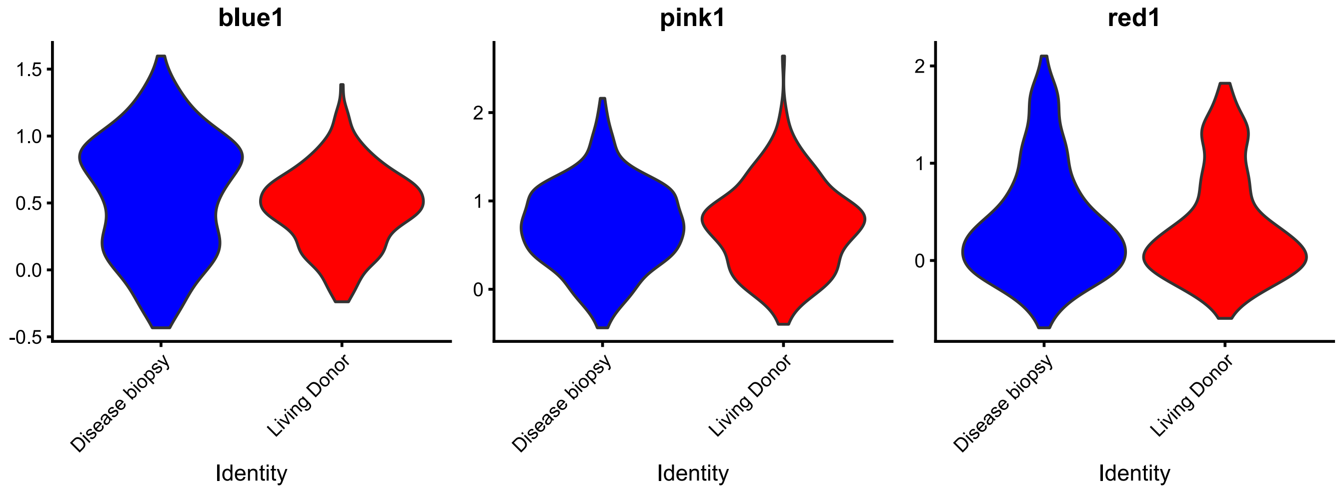

**Supplementary Figure S2. Enriched wiki pathways and regulatory potential of predicted target genes. a.** Enriched wiki pathways for the genes that were significantly differently expressed between the end and start of the afferent arteriolar cell pseudotime trajectory. **b.** Enriched wiki pathways for the genes differentially expressed between the cells at the end of trajectory in which the blue module scores was high and the cells with high pink module scores. **c.** Heatmap showing the regulatory potential of predicted target genes among the 33 module genes for the 29 bonafide ligands.

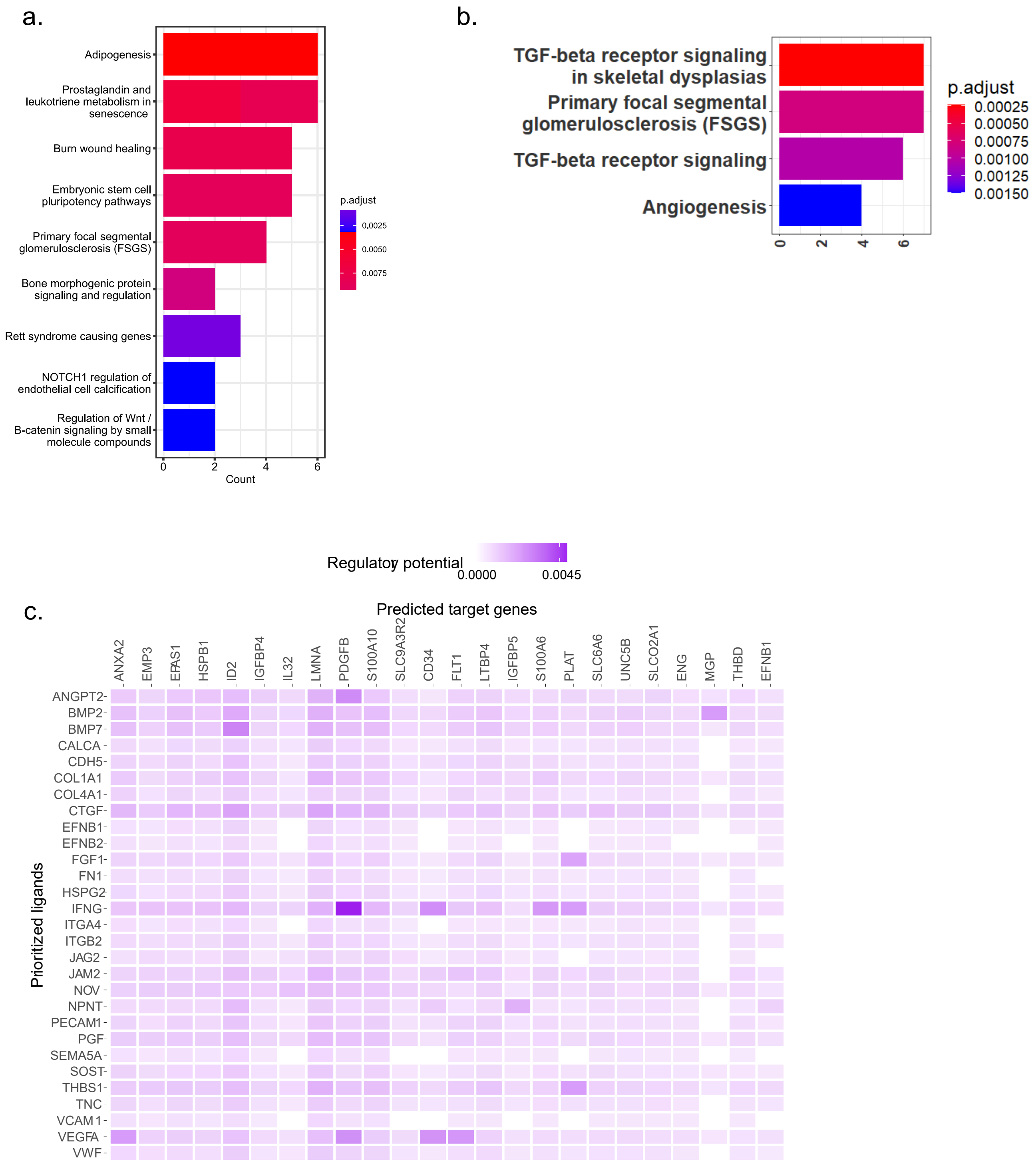

**Supplementary Figure 3** UMAP of the sixteen spatially localized niches in the 10x Visium spots based on clustering by cell type distribution.

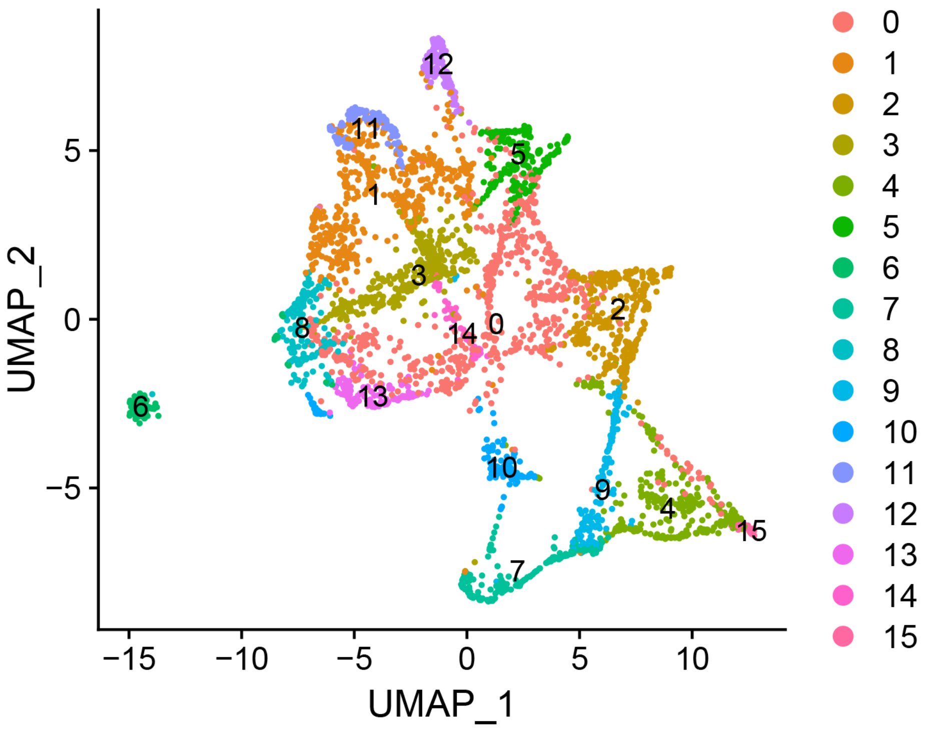

**Supplementary Figure 4. Integrated unbiased clustering analysis of six CKD visium transcriptomic datasets. a.** Heatmap showing the top 5 markers in each cluster.
**b.**Violin density plot showing the expression of afferent arteriole marker, GJ 4 in the clusters. **c.** Violin density plot showing the expression of efferent arteriole marker, S C in the clusters.

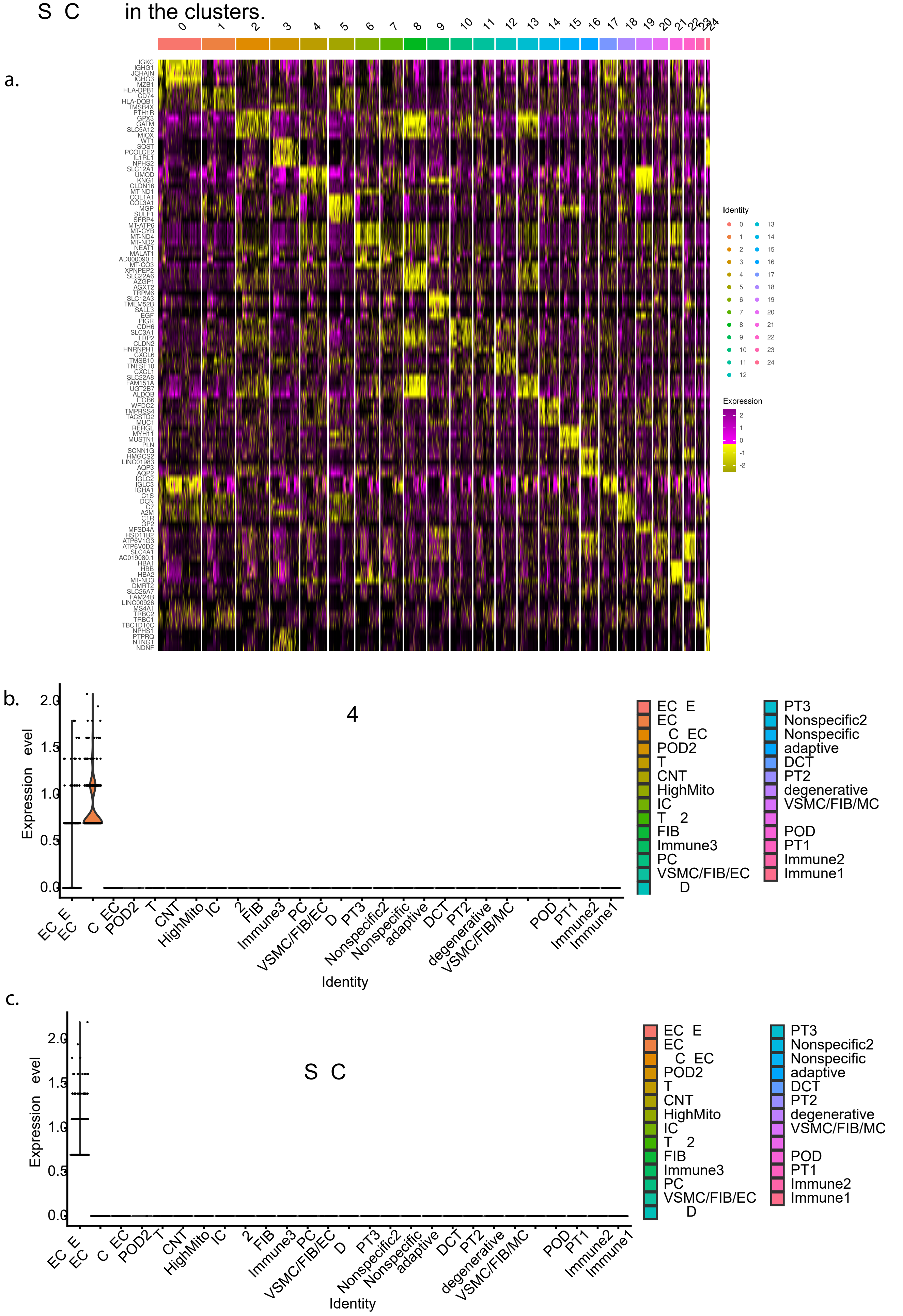

**Supplementary Table 1. Time of biopsy characteristics of participants in KPMP cohort included in this study.**

| **Total (N)** | **48** |
|---|---|
| Age (average years ± standard deviation) | 57.2 ± 15.2 |
| Sex (%) | |
| Female | 23 (48%) |
| Male | 25 (52%) |
| Race (N) | |
| Asian | 4 |
| Black or African American | 14 |
| White | 29 |
| Not reported | 1 |
| Disease category (%) | |
| Acute kidney injury (AKI) | 14 (29%) |
| Diabetic kidney disease (DKD) | 26 (54%) |
| Hypertensive chronic kidney disease (HCKD) | 8 (17%) |

**Supplementary Table 2. Time of biopsy characteristics of participants in NEPTUNE included in this study.**

| | Total N = 198 |
|---|---|
| Average age in years (SD) | 46.2 (+/-15.7) |
| Average eGFR at biopsy (SD) | 69.9 (+/-34.7) |
| Median UPCR at biopsy [IQR] | 4.0 [+/- 5.4] |
| Average % Interstitial fibrosis (SD) | 17.7 (+/-18.6) |
| Sex | |
| Female | 75 (38%) |
| Male | 123 (62%) |
| Race N (%) | |
| Asian | 21 (10%) |
| Black or African American | 51 (26%) |
| White | 132 (61%) |
| Native American/Alaskan, Pacific Islanders/Hawaiian | 1 (1%) |
| Not reported | 5 (2%) |
| Diagnosis | |
| FSGS (%) | 70 (35%) |
| APOL1 risk allele (%) | 39 (20%) |
| Immunosuppression pre- biopsy (%) | 27 (14%) |

Abbreviations: eGFR – estimated glomerular filtration rate; UPCR – urine protein creatinine ratio; FSGS – fFocal segmental glomerulosclerosis; SD – standard deviation; IQR – interquartile range.

**Supplementary Table S3. Distribution of arteriolar hyalinosis descriptor scores.** WSI of the formalin-fixed and paraffin-embedded sections from the KPMP cohort were scored for severity of arteriolar hyalinosis using a standardized semiquantitative approach: 0 (none), 1+ (mild), 2+ (moderate), or 3+ (severe)

| Arteriolar hyalinosis score | Number (Total N=45) |
|---|---|
| **No (score = 0)** | 24 |
| **Mild (score = 1)** | 13 |
| **Moderate (score = 2)** | 4 |
| **High (score = 3)** | 4 |

**Supplementary Table S4. Correlation of module genes with arteriolar hyalinosis score.** The WGCNA analysis on the meta cell expression data of the arteriolar endothelial cells yielded three gene sets/modules (blue, pink, and red) with 28, 11 and 12 genes, respectively, positively associated ($p<0.05$) with arteriolar hyalinosis

| genes | moduleColor | correlation | p value |
|---|---|---|---|
| ADGRF5 | blue | 0.1529 | 0.0132 |
| CD34 | blue | 0.1855 | 0.0026 |
| CD81 | blue | 0.2482 | 0.0000 |
| EFNB1 | blue | 0.1655 | 0.0073 |
| ENG | blue | 0.1316 | 0.0333 |
| FLT1 | blue | 0.1292 | 0.0366 |
| IGFBP4 | blue | 0.1424 | 0.0212 |
| IGFBP5 | blue | 0.1402 | 0.0233 |
| PBX1 | blue | 0.1604 | 0.0093 |
| PDGFB | blue | 0.1285 | 0.0376 |
| PRCP | blue | 0.1922 | 0.0018 |
| SLC6A6 | blue | 0.1592 | 0.0099 |
| SLC9A3R2 | blue | 0.1795 | 0.0035 |
| TCIM | blue | 0.1230 | 0.0468 |
| THBD | blue | 0.1430 | 0.0206 |
| UNC5B | blue | 0.1534 | 0.0129 |
| ANXA2 | pink | 0.2039 | 0.0009 |
| EMP3 | pink | 0.2869 | 0.0000 |
| HSPB1 | pink | 0.2356 | 0.0001 |
| IL32 | pink | 0.1789 | 0.0037 |
| LGALS1 | pink | 0.2619 | 0.0000 |
| LMNA | pink | 0.2369 | 0.0001 |
| PLAT | pink | 0.2131 | 0.0005 |
| S100A10 | pink | 0.2132 | 0.0005 |
| S100A11 | pink | 0.2373 | 0.0001 |
| S100A6 | pink | 0.3271 | 0.0000 |
| SH3BGRL3 | pink | 0.1394 | 0.0240 |
| DNAJB4 | red | 0.1963 | 0.0014 |
| EPAS1 | red | 0.1613 | 0.0089 |
| LTBP4 | red | 0.1476 | 0.0168 |
| MGP | red | 0.2025 | 0.0010 |
| PLAC9 | red | 0.1884 | 0.0022 |
| SLCO2A1 | red | 0.2253 | 0.0002 |
| ID2 | red | 0.1190 | 0.0544 |

**Supplementary Table S5. Enriched pathways along the pseudotime in afferent arteriolar cells.** Enriched wiki pathways for the differentially expressed genes at the end and start of the pseudotime trajectory in afferent arteriolar cells.

| Term | Overlap | P-value | Adjusted P-value | Odds Ratio | Combined Score | Genes |
|---|---|---|---|---|---|---|
| Adipogenesis WP236 | 6/130 | 0.0001 | 0.0160 | 9.1083 | 85.1544 | MEF2A;MBNL1;PTGIS;EPAS1;LIFR;IL6ST |
| ESC Pluripotency Pathways WP3931 | 5/116 | 0.0005 | 0.0391 | 8.4047 | 64.3515 | BMPR2;LIFR;IL6ST;SMAD6;FGF2 |
| Primary focal segmental glomerulosclerosis (FSGS) WP2572 | 26390.00 | 0.0007 | 0.0391 | 10.8966 | 79.2908 | ITGB1;JAG1;PODXL;UTRN |
| TGF-beta Signaling Pathway WP366 | 5/132 | 0.0009 | 0.0391 | 7.3399 | 51.8923 | MEF2A;ITGB1;FN1;SKIL;MET |
| Bone morphogenic protein (BMP) signaling and regulation WP1425 | 44969.00 | 0.0019 | 0.0595 | 36.4752 | 227.7372 | BMPR2;SMAD6 |
| Rett syndrome causing genes WP4312 | 17593.00 | 0.0024 | 0.0595 | 12.2494 | 73.8937 | TCF4;SHANK3;SYNE2 |
| Senescence and Autophagy in Cancer WP615 | 4/105 | 0.0028 | 0.0595 | 7.3241 | 43.0685 | IGFBP3;LAMP2;FN1;IL6ST |
| Gastrin signaling pathway WP4659 | 4/114 | 0.0038 | 0.0595 | 6.7218 | 37.5400 | ITGB1;JAG1;GNAQ;TCF4 |
| Amplification and Expansion of Oncogenic Pathways as Metastatic Traits WP3678 | 44974.00 | 0.0039 | 0.0595 | 24.3107 | 134.6502 | JAG1;EPAS1 |
| NOTCH1 regulation of endothelial cell calcification WP3413 | 44974.00 | 0.0039 | 0.0595 | 24.3107 | 134.6502 | JAG1;GJA5 |
| Regulation of Wnt/B-catenin Signaling by Small Molecule Compounds WP3664 | 44974.00 | 0.0039 | 0.0595 | 24.3107 | 134.6502 | CSNK1A1;TCF4 |
| TGF-beta receptor signaling in skeletal dysplasias WP4816 | 21245.00 | 0.0041 | 0.0595 | 10.0172 | 55.0313 | LTBP3;SMAD6;SKIL |
| Simplified Interaction Map Between LOXL4 and Oxidative Stress Pathway WP3670 | 44975.00 | 0.0044 | 0.0595 | 22.7901 | 123.6263 | FN1;EXOC6 |
| Novel intracellular components of RIG-I-like receptor (RLR) pathway WP3865 | 21976.00 | 0.0045 | 0.0595 | 9.6647 | 52.1723 | DDX17;DDX3X;CXCL12 |
| Lung fibrosis WP3624 | 23071.00 | 0.0052 | 0.0616 | 9.1801 | 48.3002 | ELN;FGF2;SKIL |
| mRNA Processing WP411 | 4/126 | 0.0054 | 0.0616 | 6.0570 | 31.6777 | HNRNPA2B1;HNRNPU;SF3B1;SRRM1 |
| Angiogenesis WP1539 | 44981.00 | 0.0078 | 0.0842 | 16.5696 | 80.4665 | TIMP2;FGF2 |
| Physiological and pathological hypertrophy of the heart WP1528 | 44982.00 | 0.0084 | 0.0858 | 15.8484 | 75.6997 | LIFR;IL6ST |
| Wnt/beta-catenin signaling pathway in leukemia WP3658 | 44983.00 | 0.0091 | 0.0858 | 15.1873 | 71.3810 | CSNK1A1;RUNX1T1 |
| Eicosanoid Synthesis WP167 | 44984.00 | 0.0098 | 0.0858 | 14.5791 | 67.4528 | PTGIS;PTGDS |

| | | | | | | |
|---|---|---|---|---|---|---|
| Regulation of Actin Cytoskeleton WP51 | 4/150 | 0.0098 | 0.0858 | 5.0552 | 23.3834 | NCKAP1;FN1;MSN;FGF2 |
| Nanoparticle-mediated activation of receptor signaling WP2643 | 44985.00 | 0.0105 | 0.0878 | 14.0176 | 63.8669 | ITGB1;FN1 |
| Eicosanoid metabolism via cyclooxygenases (COX) WP4719 | 10990.00 | 0.0120 | 0.0960 | 13.0151 | 57.5647 | PTGIS;PTGDS |
| Corticotropin-releasing hormone signaling pathway WP2355 | 34029.00 | 0.0150 | 0.1123 | 6.1108 | 25.6462 | GNAQ;ECE1;TCF4 |
| Signaling of Hepatocyte Growth Factor Receptor WP313 | 12451.00 | 0.0153 | 0.1123 | 11.3859 | 47.6264 | ITGB1;MET |
| Neovascularisation processes WP4331 | 13547.00 | 0.0179 | 0.1268 | 10.4084 | 41.8639 | JAG1;CXCL12 |
| Neural Crest Differentiation WP2064 | 3/101 | 0.0187 | 0.1275 | 5.6097 | 22.3177 | ITGB1;TCF4;FGF2 |
| Genes controlling nephrogenesis WP4823 | 15738.00 | 0.0238 | 0.1517 | 8.8825 | 33.2134 | ITGB1;CXCL12 |
| Circadian rhythm related genes WP3594 | 4/201 | 0.0258 | 0.1517 | 3.7368 | 13.6679 | DDX5;GNAQ;HNRNPU;PTGDS |
| Prostaglandin Synthesis and Regulation WP98 | 16469.00 | 0.0259 | 0.1517 | 8.4685 | 30.9470 | PTGIS;PTGDS |
| Regulation of Microtubule Cytoskeleton WP2038 | 16834.00 | 0.0270 | 0.1517 | 8.2756 | 29.9038 | DPYSL2;GNAQ |
| Focal Adhesion-PI3K-Akt-mTOR-signaling pathway WP3932 | 5/303 | 0.0270 | 0.1517 | 3.1010 | 11.2031 | ITGB1;EPAS1;FN1;FGF2;MET |
| SARS-CoV-2 altering angiogenesis via NRP1 WP5065 | 44931.00 | 0.0274 | 0.1517 | 45.1932 | 162.4933 | NRP1 |
| Osteoblast differentiation WP4787 | 3/118 | 0.0280 | 0.1517 | 4.7763 | 17.0734 | JAG1;BMPR2;FGF2 |
| VEGFA-VEGFR2 Signaling Pathway WP3888 | 6/432 | 0.0334 | 0.1728 | 2.6107 | 8.8738 | ITGB1;JAG1;IGFBP3;FN1;S1PR1;TXNIP |
| Wnt Signaling Pathway WP363 | 19025.00 | 0.0338 | 0.1728 | 7.2804 | 24.6595 | CSNK1A1;TCF4 |
| TGF-beta Receptor Signaling WP560 | 19756.00 | 0.0362 | 0.1758 | 6.9996 | 23.2251 | SMAD6;SKIL |
| Cardiac Hypertrophic Response WP2795 | 20121.00 | 0.0375 | 0.1758 | 6.8672 | 22.5558 | MEF2A;FGF2 |
| EV release from cardiac cells and their functional effects WP3297 | 44933.00 | 0.0382 | 0.1758 | 30.1258 | 98.3467 | CXCL12 |
| Glial Cell Differentiation WP2276 | 44933.00 | 0.0382 | 0.1758 | 30.1258 | 98.3467 | MSN |
| Ectoderm Differentiation WP2858 | 3/138 | 0.0416 | 0.1867 | 4.0646 | 12.9246 | PTPRB;PODXL;SKIL |
| Mesodermal commitment pathway WP2857 | 3/147 | 0.0486 | 0.2129 | 3.8088 | 11.5185 | BMPR2;TCF4;SMAD6 |
| Oncostatin M Signaling Pathway WP2374 | 23774.00 | 0.0506 | 0.2167 | 5.7743 | 17.2253 | LIFR;IL6ST |
| Mammary gland development pathway - Involution (Stage 4 of 4) WP2815 | 44936.00 | 0.0541 | 0.2195 | 20.0808 | 58.5571 | IL6ST |
| SARS-CoV-2 and COVID-19 Pathway WP4846 | 44936.00 | 0.0541 | 0.2195 | 20.0808 | 58.5571 | NRP1 |

| | | | | | | |
|---|---|---|---|---|---|---|
| RAC1/PAK1/p38/MMP2 Pathway WP3303 | 24869.00 | 0.0549 | 0.2195 | 5.5110 | 15.9972 | ITGB1;FN1 |
| Myometrial relaxation and contraction pathways WP289 | 3/156 | 0.0561 | 0.2198 | 3.5832 | 10.3189 | CALD1;IGFBP3;GNAQ |
| SRF and miRs in Smooth Muscle Differentiation and Proliferation WP1991 | 44937.00 | 0.0594 | 0.2277 | 18.0718 | 51.0257 | MEF2A |
| 16p11.2 proximal deletion syndrome WP4949 | 27061.00 | 0.0637 | 0.2336 | 5.0502 | 13.9065 | IGFBP3;MSN |
| Mammary gland development pathway - Puberty (Stage 2 of 4) WP2814 | 44939.00 | 0.0698 | 0.2336 | 15.0583 | 40.0837 | FN1 |
| Cell Differentiation - Index WP2029 | 44939.00 | 0.0698 | 0.2336 | 15.0583 | 40.0837 | MEF2A |
| Cell-type Dependent Selectivity of CCK2R Signaling WP3679 | 44939.00 | 0.0698 | 0.2336 | 15.0583 | 40.0837 | GNAQ |
| Development of pulmonary dendritic cells and macrophage subsets WP3892 | 44939.00 | 0.0698 | 0.2336 | 15.0583 | 40.0837 | TCF4 |
| ncRNAs involved in STAT3 signaling in hepatocellular carcinoma WP4337 | 44939.00 | 0.0698 | 0.2336 | 15.0583 | 40.0837 | IL6ST |
| Regulatory circuits of the STAT3 signaling pathway WP4538 | 28522.00 | 0.0698 | 0.2336 | 4.7834 | 12.7324 | LIFR;IL6ST |
| Somatic sex determination WP4814 | 44940.00 | 0.0750 | 0.2417 | 13.8993 | 36.0063 | PTGDS |
| H19 action Rb-E2F1 signaling and CDK-Beta-catenin activity WP3969 | 44940.00 | 0.0750 | 0.2417 | 13.8993 | 36.0063 | JAG1 |
| MicroRNAs in cardiomyocyte hypertrophy WP1544 | 30713.00 | 0.0794 | 0.2417 | 4.4321 | 11.2297 | IL6ST;FGF2 |
| EGFR Tyrosine Kinase Inhibitor Resistance WP4806 | 30713.00 | 0.0794 | 0.2417 | 4.4321 | 11.2297 | FGF2;MET |
| Mammary gland development pathway - Embryonic development (Stage 1 of 4) WP2813 | 44941.00 | 0.0801 | 0.2417 | 12.9058 | 32.5775 | ITGB1 |
| FOXP3 in COVID-19 WP5063 | 44941.00 | 0.0801 | 0.2417 | 12.9058 | 32.5775 | IL6ST |
| Airway smooth muscle cell contraction WP4962 | 44943.00 | 0.0903 | 0.2420 | 11.2915 | 27.1505 | GNAQ |
| Canonical and non-canonical TGF-B signaling WP3874 | 44943.00 | 0.0903 | 0.2420 | 11.2915 | 27.1505 | BMPR2 |
| Cells and molecules involved in local acute inflammatory response WP4493 | 44943.00 | 0.0903 | 0.2420 | 11.2915 | 27.1505 | ITGB1 |
| miR-509-3p alteration of YAP1/ECM axis WP3967 | 44943.00 | 0.0903 | 0.2420 | 11.2915 | 27.1505 | FN1 |
| Nephrogenesis WP5052 | 44943.00 | 0.0903 | 0.2420 | 11.2915 | 27.1505 | JAG1 |
| GPCRs, Other WP117 | 33270.00 | 0.0910 | 0.2420 | 4.0821 | 9.7857 | ADGRF5;S1PR1 |
| TGF-B Signaling in Thyroid Cells for Epithelial-Mesenchymal Transition WP3859 | 44944.00 | 0.0954 | 0.2420 | 10.6267 | 24.9736 | FN1 |
| Transcription co-factors SKI and SKIL protein partners WP4533 | 44944.00 | 0.0954 | 0.2420 | 10.6267 | 24.9736 | SKIL |

| | | | | | | |
|---|---|---|---|---|---|---|
| MFAP5 effect on permeability and motility of endothelial cells via cytoskeleton rearrangement WP4560 | 44944.00 | 0.0954 | 0.2420 | 10.6267 | 24.9736 | LPP |
| LncRNA involvement in canonical Wnt signaling and colorectal cancer WP4258 | 34366.00 | 0.0961 | 0.2420 | 3.9483 | 9.2490 | CSNK1A1;HNRNPU |
| Focal Adhesion WP306 | 3/198 | 0.0979 | 0.2420 | 2.8054 | 6.5179 | ITGB1;FN1;MET |
| Small cell lung cancer WP4658 | 35096.00 | 0.0995 | 0.2420 | 3.8639 | 8.9148 | ITGB1;FN1 |
| Small Ligand GPCRs WP247 | 44945.00 | 0.1004 | 0.2420 | 10.0359 | 23.0696 | S1PR1 |
| Cell Differentiation - Index expanded WP2023 | 44945.00 | 0.1004 | 0.2420 | 10.0359 | 23.0696 | MEF2A |
| Overview of nanoparticle effects WP3287 | 44945.00 | 0.1004 | 0.2420 | 10.0359 | 23.0696 | FN1 |
| Complement system WP2806 | 35462.00 | 0.1013 | 0.2420 | 3.8231 | 8.7542 | CD93;CSNK1A1 |
| Pathways Regulating Hippo Signaling WP4540 | 35827.00 | 0.1030 | 0.2431 | 3.7831 | 8.5977 | GNAQ;MET |
| Serotonin Receptor 2 and ELK-SRF/GATA4 signaling WP732 | 44946.00 | 0.1054 | 0.2455 | 9.5072 | 21.3924 | GNAQ |
| 7q11.23 copy number variation syndrome WP4932 | 2/104 | 0.1137 | 0.2587 | 3.5595 | 7.7388 | ELN;SF3B1 |
| miRNA targets in ECM and membrane receptors WP2911 | 44948.00 | 0.1153 | 0.2587 | 8.6009 | 18.5799 | FN1 |
| PKC-gamma calcium signaling pathway in ataxia WP4760 | 44948.00 | 0.1153 | 0.2587 | 8.6009 | 18.5799 | GNAQ |
| Cancer immunotherapy by PD-1 blockade WP4585 | 44949.00 | 0.1202 | 0.2587 | 8.2095 | 17.3917 | NFAT5 |
| Glutathione metabolism WP100 | 44949.00 | 0.1202 | 0.2587 | 8.2095 | 17.3917 | GPX3 |
| Hippo-Yap signaling pathway WP4537 | 44949.00 | 0.1202 | 0.2587 | 8.2095 | 17.3917 | STK38L |
| PI3K-Akt signaling pathway WP4172 | 4/340 | 0.1209 | 0.2587 | 2.1755 | 4.5959 | ITGB1;FN1;FGF2;MET |
| IL1 and megakaryocytes in obesity WP2865 | 44950.00 | 0.1251 | 0.2616 | 7.8522 | 16.3217 | TIMP2 |
| Unfolded protein response WP4925 | 44950.00 | 0.1251 | 0.2616 | 7.8522 | 16.3217 | TXNIP |
| Signal Transduction of S1P Receptor WP26 | 44951.00 | 0.1300 | 0.2657 | 7.5246 | 15.3540 | S1PR1 |
| Mammalian disorder of sexual development WP4842 | 44951.00 | 0.1300 | 0.2657 | 7.5246 | 15.3540 | PTGDS |
| 1q21.1 copy number variation syndrome WP4905 | 44953.00 | 0.1396 | 0.2792 | 6.9451 | 13.6744 | GJA5 |
| Canonical and non-canonical Notch signaling WP3845 | 44953.00 | 0.1396 | 0.2792 | 6.9451 | 13.6744 | JAG1 |
| Hippo-Merlin Signaling Dysregulation WP4541 | 2/120 | 0.1434 | 0.2803 | 3.0743 | 5.9706 | ITGB1;MET |
| TLR4 Signaling and Tolerance WP3851 | 44954.00 | 0.1444 | 0.2803 | 6.6875 | 12.9421 | TRAM1 |
| T-Cell Receptor and Co-stimulatory Signaling WP2583 | 44955.00 | 0.1491 | 0.2803 | 6.4484 | 12.2703 | CSNK1A1 |

| | | | | | | |
|---|---|---|---|---|---|---|
| Inflammatory Response Pathway WP453 | 44956.00 | 0.1539 | 0.2803 | 6.2257 | 11.6522 | FN1 |
| Sphingolipid pathway WP1422 | 44956.00 | 0.1539 | 0.2803 | 6.2257 | 11.6522 | SERINC1 |
| Matrix Metalloproteinases WP129 | 44956.00 | 0.1539 | 0.2803 | 6.2257 | 11.6522 | TIMP2 |
| Cell migration and invasion through p75NTR WP4561 | 44956.00 | 0.1539 | 0.2803 | 6.2257 | 11.6522 | EFNB2 |
| Oligodendrocyte specification and differentiation, leading to myelin components for CNS WP4304 | 44956.00 | 0.1539 | 0.2803 | 6.2257 | 11.6522 | FGF2 |
| Extracellular vesicle-mediated signaling in recipient cells WP2870 | 44956.00 | 0.1539 | 0.2803 | 6.2257 | 11.6522 | MET |
| Gastric Cancer Network 2 WP2363 | 44957.00 | 0.1586 | 0.2861 | 6.0179 | 11.0821 | RNF144B |
| White fat cell differentiation WP4149 | 11689.00 | 0.1633 | 0.2861 | 5.8235 | 10.5548 | MECOM |
| Ovarian infertility WP34 | 11689.00 | 0.1633 | 0.2861 | 5.8235 | 10.5548 | SYNE2 |
| Hair Follicle Development: Organogenesis - Part 2 of 3 WP2839 | 11689.00 | 0.1633 | 0.2861 | 5.8235 | 10.5548 | ITGB1 |
| Endothelin Pathways WP2197 | 12055.00 | 0.1679 | 0.2887 | 5.6412 | 10.0659 | ECE1 |
| Oxidative Stress WP408 | 12055.00 | 0.1679 | 0.2887 | 5.6412 | 10.0659 | GPX3 |
| Type 2 papillary renal cell carcinoma WP4241 | 12420.00 | 0.1725 | 0.2939 | 5.4700 | 9.6118 | EPAS1 |
| The influence of laminopathies on Wnt signaling WP4844 | 12785.00 | 0.1771 | 0.2963 | 5.3088 | 9.1889 | CSNK1A1 |
| Overview of leukocyte-intrinsic Hippo pathway functions WP4542 | 12785.00 | 0.1771 | 0.2963 | 5.3088 | 9.1889 | STK38L |
| Melatonin metabolism and effects WP3298 | 13516.00 | 0.1863 | 0.3033 | 5.0134 | 8.4257 | ECE1 |
| Factors and pathways affecting insulin-like growth factor (IGF1)-Akt signaling WP3850 | 13516.00 | 0.1863 | 0.3033 | 5.0134 | 8.4257 | ITGB1 |
| Photodynamic therapy-induced HIF-1 survival signaling WP3614 | 13516.00 | 0.1863 | 0.3033 | 5.0134 | 8.4257 | IGFBP3 |
| Brain-derived neurotrophic factor (BDNF) signaling pathway WP2380 | 2/144 | 0.1904 | 0.3052 | 2.5516 | 4.2316 | MEF2A;DPYSL2 |
| Sleep regulation WP3591 | 13881.00 | 0.1908 | 0.3052 | 4.8776 | 8.0805 | PTGDS |
| Male infertility WP4673 | 2/146 | 0.1945 | 0.3085 | 2.5159 | 4.1199 | CDC42BPA;CLU |
| Calcium Regulation in the Cardiac Cell WP536 | 2/150 | 0.2025 | 0.3164 | 2.4474 | 3.9083 | GJA5;GNAQ |
| Integrated breast cancer pathway WP1984 | 2/152 | 0.2066 | 0.3164 | 2.4146 | 3.8080 | BMPR2;SMAD6 |
| Hepatitis B infection WP4666 | 2/152 | 0.2066 | 0.3164 | 2.4146 | 3.8080 | DDX3X;HSPG2 |
| Fas ligand pathway and stress induction of heat shock proteins WP314 | 15342.00 | 0.2086 | 0.3164 | 4.4009 | 6.8970 | SPTAN1 |
| Breast cancer pathway WP4262 | 2/154 | 0.2106 | 0.3164 | 2.3825 | 3.7112 | CSNK1A1;FGF2 |
| IL-6 signaling pathway WP364 | 15707.00 | 0.2130 | 0.3164 | 4.2959 | 6.6428 | IL6ST |
| Interleukin-11 Signaling Pathway WP2332 | 16072.00 | 0.2174 | 0.3164 | 4.1958 | 6.4027 | IL6ST |
| Heart Development WP1591 | 16072.00 | 0.2174 | 0.3164 | 4.1958 | 6.4027 | BMPR2 |

| | | | | | | |
|---|---|---|---|---|---|---|
| Hedgehog Signaling Pathway WP4249 | 16072.00 | 0.2174 | 0.3164 | 4.1958 | 6.4027 | CSNK1A1 |
| Renin-angiotensin-aldosterone system (RAAS) WP4756 | 16072.00 | 0.2174 | 0.3164 | 4.1958 | 6.4027 | GNAQ |
| Nephrotic syndrome WP4758 | 16438.00 | 0.2218 | 0.3164 | 4.1002 | 6.1755 | PODXL |
| Notch Signaling WP268 | 16438.00 | 0.2218 | 0.3164 | 4.1002 | 6.1755 | JAG1 |
| Epithelial to mesenchymal transition in colorectal cancer WP4239 | 2/160 | 0.2228 | 0.3164 | 2.2914 | 3.4401 | JAG1;FN1 |
| Envelope proteins and their potential roles in EDMD physiopathology WP4535 | 16803.00 | 0.2261 | 0.3164 | 4.0089 | 5.9605 | SYNE2 |
| Mechanoregulation and pathology of YAP/TAZ via Hippo and non-Hippo mechanisms WP4534 | 17168.00 | 0.2304 | 0.3164 | 3.9215 | 5.7567 | ITGB1 |
| Development of ureteric collection system WP5053 | 17168.00 | 0.2304 | 0.3164 | 3.9215 | 5.7567 | ITGB1 |
| NO/cGMP/PKG mediated Neuroprotection WP4008 | 17168.00 | 0.2304 | 0.3164 | 3.9215 | 5.7567 | PDE3A |
| Energy Metabolism WP1541 | 17168.00 | 0.2304 | 0.3164 | 3.9215 | 5.7567 | MEF2A |
| Differentiation Pathway WP2848 | 17533.00 | 0.2347 | 0.3175 | 3.8379 | 5.5632 | FGF2 |
| Exercise-induced Circadian Regulation WP410 | 17533.00 | 0.2347 | 0.3175 | 3.8379 | 5.5632 | NCKAP1 |
| Hepatitis C and Hepatocellular Carcinoma WP3646 | 17899.00 | 0.2389 | 0.3209 | 3.7578 | 5.3795 | PODXL |
| Synaptic signaling pathways associated with autism spectrum disorder WP4539 | 18264.00 | 0.2432 | 0.3242 | 3.6809 | 5.2048 | SHANK3 |
| Copper homeostasis WP3286 | 18994.00 | 0.2516 | 0.3306 | 3.5362 | 4.8801 | XAF1 |
| One-carbon metabolism and related pathways WP3940 | 18994.00 | 0.2516 | 0.3306 | 3.5362 | 4.8801 | GPX3 |
| Cardiac Progenitor Differentiation WP2406 | 19360.00 | 0.2557 | 0.3314 | 3.4680 | 4.7291 | FGF2 |
| Phosphodiesterases in neuronal function WP4222 | 19360.00 | 0.2557 | 0.3314 | 3.4680 | 4.7291 | PDE3A |
| The Overlap Between Signal Transduction Pathways that Contribute to a Range of LMNA Laminopathies WP4879 | 20090.00 | 0.2640 | 0.3350 | 3.3392 | 4.4473 | CSNK1A1 |
| Pathogenic Escherichia coli infection WP2272 | 20090.00 | 0.2640 | 0.3350 | 3.3392 | 4.4473 | ITGB1 |
| Hematopoietic Stem Cell Differentiation WP2849 | 20090.00 | 0.2640 | 0.3350 | 3.3392 | 4.4473 | CD34 |
| Vitamin D Receptor Pathway WP2877 | 2/182 | 0.2680 | 0.3377 | 2.0091 | 2.6457 | IGFBP3;TIMP2 |
| Ras signaling WP4223 | 2/184 | 0.2721 | 0.3406 | 1.9868 | 2.5861 | RAPGEF5;MET |
| Complement and Coagulation Cascades WP558 | 21186.00 | 0.2762 | 0.3434 | 3.1630 | 4.0695 | CLU |
| MET in type 1 papillary renal cell carcinoma WP4205 | 21551.00 | 0.2802 | 0.3461 | 3.1083 | 3.9541 | MET |

| | | | | | | |
|---|---|---|---|---|---|---|
| Notch Signaling Pathway Netpath WP61 | 22282.00 | 0.2882 | 0.3536 | 3.0044 | 3.7374 | JAG1 |
| Endometrial cancer WP4155 | 23012.00 | 0.2961 | 0.3608 | 2.9072 | 3.5379 | FGF2 |
| Endochondral Ossification with Skeletal Dysplasias WP4808 | 23377.00 | 0.3001 | 0.3608 | 2.8609 | 3.4439 | FGF2 |
| Endochondral Ossification WP474 | 23377.00 | 0.3001 | 0.3608 | 2.8609 | 3.4439 | FGF2 |
| AGE/RAGE pathway WP2324 | 24108.00 | 0.3078 | 0.3608 | 2.7726 | 3.2667 | MSN |
| Association Between Physico-Chemical Features and Toxicity Associated Pathways WP3680 | 24108.00 | 0.3078 | 0.3608 | 2.7726 | 3.2667 | FN1 |
| Thyroid stimulating hormone (TSH) signaling pathway WP2032 | 24108.00 | 0.3078 | 0.3608 | 2.7726 | 3.2667 | GNAQ |
| SARS-CoV-2 innate immunity evasion and cell-specific immune response WP5039 | 24108.00 | 0.3078 | 0.3608 | 2.7726 | 3.2667 | CXCL12 |
| Folate Metabolism WP176 | 25204.00 | 0.3193 | 0.3719 | 2.6499 | 3.0249 | GPX3 |
| MECP2 and Associated Rett Syndrome WP3584 | 26299.00 | 0.3306 | 0.3826 | 2.5375 | 2.8084 | FGF2 |
| Chromosomal and microsatellite instability in colorectal cancer WP4216 | 26665.00 | 0.3344 | 0.3840 | 2.5021 | 2.7412 | CSNK1A1 |
| Ciliary landscape WP4352 | 2/216 | 0.3375 | 0.3840 | 1.6870 | 1.8323 | DDX5;EXOC6 |
| Arrhythmogenic Right Ventricular Cardiomyopathy WP2118 | 27030.00 | 0.3381 | 0.3840 | 2.4677 | 2.6763 | ITGB1 |
| Prolactin Signaling Pathway WP2037 | 27760.00 | 0.3454 | 0.3899 | 2.4017 | 2.5529 | ITGB1 |
| Glioblastoma signaling pathways WP2261 | 29952.00 | 0.3670 | 0.4118 | 2.2231 | 2.2284 | MET |
| Alzheimer's disease WP2059 | 30317.00 | 0.3705 | 0.4132 | 2.1959 | 2.1801 | GNAQ |
| Pathways in clear cell renal cell carcinoma WP4018 | 31048.00 | 0.3775 | 0.4185 | 2.1434 | 2.0879 | RAPGEF5 |
| ncRNAs involved in Wnt signaling in hepatocellular carcinoma WP4336 | 31413.00 | 0.3810 | 0.4198 | 2.1181 | 2.0438 | CSNK1A1 |
| Selenium Micronutrient Network WP15 | 32509.00 | 0.3913 | 0.4260 | 2.0456 | 1.9193 | GPX3 |
| Allograft Rejection WP2328 | 32509.00 | 0.3913 | 0.4260 | 2.0456 | 1.9193 | CXCL12 |
| G Protein Signaling Pathways WP35 | 33970.00 | 0.4048 | 0.4381 | 1.9562 | 1.7693 | GNAQ |
| Integrin-mediated Cell Adhesion WP185 | 1/101 | 0.4308 | 0.4636 | 1.7990 | 1.5150 | ITGB1 |
| IL-18 signaling pathway WP4754 | 2/272 | 0.4470 | 0.4782 | 1.3333 | 1.0735 | MEF2A;FN1 |
| Wnt signaling WP428 | 1/115 | 0.4737 | 0.5038 | 1.5770 | 1.1783 | CSNK1A1 |
| Spinal Cord Injury WP2431 | 1/118 | 0.4825 | 0.5102 | 1.5363 | 1.1198 | EFNB2 |
| Ebola Virus Pathway on Host WP4217 | 1/129 | 0.5134 | 0.5398 | 1.4035 | 0.9358 | ITGB1 |
| Regulation of toll-like receptor signaling pathway WP1449 | 1/139 | 0.5399 | 0.5644 | 1.3011 | 0.8020 | SMAD6 |
| Endoderm differentiation WP2853 | 1/141 | 0.5450 | 0.5666 | 1.2824 | 0.7783 | TCF4 |
| NRF2 pathway WP2884 | 1/146 | 0.5576 | 0.5764 | 1.2379 | 0.7231 | GPX3 |

| | | | | | | |
|---|---|---|---|---|---|---|
| Sudden Infant Death Syndrome (SIDS) Susceptibility Pathways WP706 | 1/158 | 0.5864 | 0.6028 | 1.1426 | 0.6099 | ECE1 |
| EGF/EGFR signaling pathway WP437 | 1/162 | 0.5956 | 0.6088 | 1.1139 | 0.5773 | MEF2A |
| Chemokine signaling pathway WP3929 | 1/164 | 0.6001 | 0.6100 | 1.1002 | 0.5619 | CXCL12 |
| Metapathway biotransformation Phase I and II WP702 | 1/183 | 0.6405 | 0.6476 | 0.9844 | 0.4385 | GPX3 |
| MAPK Signaling Pathway WP382 | 1/246 | 0.7478 | 0.7519 | 0.7289 | 0.2118 | FGF2 |
| Nuclear Receptors Meta-Pathway WP2882 | 1/319 | 0.8330 | 0.8330 | 0.5595 | 0.1022 | GPX3 |

**Supplementary Table S6. Pathway differences in efferent cells with high blue and pink module scores.** Enriched wiki pathways for the differentially expressed genes between efferent arteriolar cells with high blue module scores and high pink module scores.

| ID | Description | GeneRatio | Bg Ratio | pvalue | p.adjust | qvalue | geneID | Count | |
|---|---|---|---|---|---|---|---|---|---|
| WP4816 | WP4816 | TGF-beta receptor signaling in skeletal dysplasias | 7/172 | 59/8085 | 0.0002 | 0.0002 | 0.061951529 | 6497/7048/6498/4054/4093/4091/2033 | 7 |
| WP2572 | WP2572 | Primary focal segmental glomerulosclerosis | 7/172 | 72/8085 | 0.0008 | 0.0008 | 0.091453934 | 950/23607/5747/7099/4040/64423/5335 | 7 |
| WP560 | WP560 | TGF-beta receptor signaling | 6/172 | 55/8085 | 0.0010 | 0.0010 | 0.091453934 | 6497/7048/6498/4093/4091/2033 | 6 |
| WP1539 | WP1539 | Angiogenesis | 4/172 | 24/8085 | 0.0015 | 0.0015 | 0.099520346 | 3791/5747/2321/5335 | 4 |
| WP313 | WP313 | Hepatocyte growth factor receptor signaling | 4/172 | 34/8085 | 0.0056 | 0.0056 | 0.220889754 | 3672/5747/2889/5829 | 4 |
| WP5144 | WP5144 | NRP1-triggered signaling pathways in pancreatic cancer | 5/172 | 54/8085 | 0.0056 | 0.0056 | 0.220889754 | 7048/3791/1105/5747/2321 | 5 |
| WP411 | WP411 | mRNA processing | 8/172 | 128/8085 | 0.0059 | 0.0059 | 0.220889754 | 6421/10181/8899/10658/23405/2521/6431/5725 | 8 |
| WP366 | WP366 | TGF-beta signaling pathway | 8/172 | 133/8085 | 0.0073 | 0.0073 | 0.24228658 | 6497/7048/6498/9765/7071/5747/64750/2033 | 8 |
| WP306 | WP306 | Focal adhesion | 10/172 | 198/8085 | 0.0095 | 0.0095 | 0.268837463 | 8503/10188/3791/3672/5747/2889/80310/3678/5829/2321 | 10 |
| WP3888 | WP3888 | VEGFA-VEGFR2 signaling | 17/172 | 436/8085 | 0.0112 | 0.0112 | 0.268837463 | 10628/3791/253260/4646/2887/5747/2889/51564/91010/5829/2321/8650/22899/5335/23476/2130/9510 | 17 |
| WP2369 | WP2369 | Histone modifications | 5/172 | 65/8085 | 0.0121 | 0.0121 | 0.268837463 | 55870/64324/58508/51111/4297 | 5 |
| WP3931 | WP3931 | Embryonic stem cell pluripotency pathways | 7/172 | 117/8085 | 0.0122 | 0.0122 | 0.268837463 | 10097/659/3977/324/4040/4093/4091 | 7 |
| WP3658 | WP3658 | Wnt/beta-catenin signaling pathway in leukemia | 3/172 | 26/8085 | 0.0172 | 0.0172 | 0.330291602 | 324/4040/3728 | 3 |
| WP2643 | WP2643 | Nanoparticle-mediated activation of receptor signaling | 3/172 | 28/8085 | 0.0210 | 0.0210 | 0.330291602 | 3672/5747/5829 | 3 |

| | | | | | | | | | |
|---|---|---|---|---|---|---|---|---|---|
| WP4217 | WP4217 | Ebola virus infection in host | 7/172 | 131/8085 | 0.0215 | 0.0215 | 0.330291602 | 8503/5610/3672/7099/3678/2621/2033 | 7 |
| WP437 | WP437 | EGF/EGFR signaling pathway | 8/172 | 162/8085 | 0.0221 | 0.0221 | 0.330291602 | 9922/10188/253260/2887/6197/5747/9101/5335 | 8 |
| WP236 | WP236 | Adipogenesis | 7/172 | 132/8085 | 0.0223 | 0.0223 | 0.330291602 | 2034/8609/25937/4154/3977/196/10658 | 7 |
| WP4747 | WP4747 | Netrin-UNC5B signaling pathway | 4/172 | 52/8085 | 0.0243 | 0.0243 | 0.330291602 | 249/3791/5747/23365 | 4 |
| WP3844 | WP3844 | PI3K-AKT-mTOR signaling pathway and therapeutic opportunities | 3/172 | 30/8085 | 0.0252 | 0.0252 | 0.330291602 | 8503/253260/2887 | 3 |
| WP2864 | WP2864 | Apoptosis-related network due to altered Notch3 in ovarian cancer | 4/172 | 53/8085 | 0.0258 | 0.0258 | 0.330291602 | 5747/3309/8065/5585 | 4 |
| WP4868 | WP4868 | Type I interferon induction and signaling during SARS-CoV-2 infection | 3/172 | 31/8085 | 0.0275 | 0.0275 | 0.330291602 | 5610/7099/10379 | 3 |
| WP3594 | WP3594 | Circadian rhythm genes | 9/172 | 201/8085 | 0.0275 | 0.0275 | 0.330291602 | 6421/9967/196/7071/23064/4297/5927/463/2033 | 9 |
| WP4172 | WP4172 | PI3K-Akt signaling pathway | 13/172 | 340/8085 | 0.0291 | 0.0291 | 0.331650568 | 8503/5586/1435/3791/3672/5747/7099/80310/3678/2321/10161/3643/5585 | 13 |
| WP3995 | WP3995 | Prion disease pathway | 3/172 | 33/8085 | 0.0324 | 0.0324 | 0.331650568 | 5747/3309/2033 | 3 |
| WP4806 | WP4806 | EGFR tyrosine kinase inhibitor resistance | 5/172 | 84/8085 | 0.0330 | 0.0330 | 0.331650568 | 8503/3791/80310/2621/5335 | 5 |
| WP4241 | WP4241 | Type 2 papillary renal cell carcinoma | 3/172 | 34/8085 | 0.0350 | 0.0350 | 0.331650568 | 6421/2034/2033 | 3 |
| WP4659 | WP4659 | Gastrin signaling pathway | 6/172 | 115/8085 | 0.0357 | 0.0357 | 0.331650568 | 8503/5747/51564/5829/5583/5335 | 6 |
| WP4018 | WP4018 | Clear cell renal cell carcinoma pathways | 5/172 | 86/8085 | 0.0360 | 0.0360 | 0.331650568 | 55193/3791/2887/2321/2033 | 5 |
| WP304 | WP304 | Kit receptor signaling pathway | 4/172 | 59/8085 | 0.0364 | 0.0364 | 0.331650568 | 2887/6197/5335/2033 | 4 |
| WP61 | WP61 | Notch signaling pathway | 4/172 | 61/8085 | 0.0404 | 0.0404 | 0.332163148 | 23013/84441/8650/2033 | 4 |

| | | | | | | | | | |
|---|---|---|---|---|---|---|---|---|---|
| WP4787 | WP4787 | Osteoblast differentiation and related diseases | 6/172 | 119/8085 | 0.0411 | 0.0411 | 0.332163148 | 8503/659/5286/4040/4093/5583 | 6 |
| WP51 | WP51 | Regulation of actin cytoskeleton | 7/172 | 152/8085 | 0.0432 | 0.0432 | 0.332163148 | 8503/1131/3672/324/5747/5286/5829 | 7 |
| WP4844 | WP4844 | Influence of laminopathies on Wnt signaling | 3/172 | 37/8085 | 0.0434 | 0.0434 | 0.332163148 | 26092/324/23405 | 3 |
| WP619 | WP619 | Type II interferon signaling | 3/172 | 37/8085 | 0.0434 | 0.0434 | 0.332163148 | 5610/10379/4261 | 3 |
| WP4541 | WP4541 | Hippo-Merlin signaling dysregulation | 6/172 | 121/8085 | 0.0440 | 0.0440 | 0.332163148 | 3791/3672/5747/3678/2321/3643 | 6 |
| WP2374 | WP2374 | Oncostatin M signaling pathway | 4/172 | 65/8085 | 0.0492 | 0.0492 | 0.351074243 | 3977/253260/5829/5583 | 4 |
| WP3651 | WP3651 | Pathways affected in adenoid cystic carcinoma | 4/172 | 65/8085 | 0.0492 | 0.0492 | 0.351074243 | 55914/64324/58508/2033 | 4 |
| WP3680 | WP3680 | Physico-chemical features and toxicity-associated pathways | 4/172 | 66/8085 | 0.0515 | 0.0515 | 0.358093124 | 10097/324/5747/5335 | 4 |
| WP481 | WP481 | Insulin signaling | 7/172 | 160/8085 | 0.0543 | 0.0543 | 0.367779335 | 8503/2887/6197/2889/5286/5583/3643 | 7 |
| WP4540 | WP4540 | Hippo signaling regulation pathways | 5/172 | 98/8085 | 0.0575 | 0.0575 | 0.379677165 | 23683/3791/2321/5583/3643 | 5 |
| WP3915 | WP3915 | Angiopoietin-like protein 8 regulatory pathway | 6/172 | 132/8085 | 0.0622 | 0.0622 | 0.400716547 | 8503/253260/6197/2889/5286/3643 | 6 |
| WP185 | WP185 | Integrin-mediated cell adhesion | 5/172 | 102/8085 | 0.0660 | 0.0660 | 0.411739879 | 3672/5747/2889/3678/5829 | 5 |
| WP3584 | WP3584 | MECP2 and associated Rett syndrome | 4/172 | 73/8085 | 0.0695 | 0.0695 | 0.411739879 | 23435/5978/4204/2521 | 4 |
| WP4300 | WP4300 | Extracellular vesicles in the crosstalk of cardiac cells | 2/172 | 21/8085 | 0.0726 | 0.0726 | 0.411739879 | 3791/7099 | 2 |
| WP4535 | WP4535 | Envelope proteins and their potential roles in EDMD physiopathology | 3/172 | 46/8085 | 0.0739 | 0.0739 | 0.411739879 | 23345/196883/23224 | 3 |
| WP4861 | WP4861 | Endoplasmic reticulum stress response in coronavirus infection | 3/172 | 46/8085 | 0.0739 | 0.0739 | 0.411739879 | 5610/5514/3309 | 3 |
| WP2586 | WP2586 | Aryl hydrocarbon receptor pathway | 3/172 | 47/8085 | 0.0777 | 0.0777 | 0.411739879 | 135112/196/2033 | 3 |

| | | | | | | | | | |
|---|---|---|---|---|---|---|---|---|---|
| WP4312 | WP4312 | Rett syndrome causing genes | 3/172 | 48/8085 | 0.0817 | 0.0817 | 0.411739879 | 4204/23224/85358 | 3 |
| WP4725 | WP4725 | Sphingolipid metabolism overview | 2/172 | 23/8085 | 0.0851 | 0.0851 | 0.411739879 | 8613/10558 | 2 |
| WP4925 | WP4925 | Unfolded protein response | 2/172 | 23/8085 | 0.0851 | 0.0851 | 0.411739879 | 10628/3309 | 2 |
| WP5044 | WP5044 | Kynurenine pathway and links to cell senescence | 2/172 | 23/8085 | 0.0851 | 0.0851 | 0.411739879 | 196/7099 | 2 |
| WP5181 | WP5181 | Roles of ceramides in development of insulin resistance | 2/172 | 23/8085 | 0.0851 | 0.0851 | 0.411739879 | 5610/3643 | 2 |
| WP5087 | WP5087 | Malignant pleural mesothelioma | 14/172 | 440/8085 | 0.0853 | 0.0853 | 0.411739879 | 5928/1435/25937/3791/3672/324/58508/6197/5747/80310/4040/2321/3643/9510 | 14 |
| WP4758 | WP4758 | Nephrotic syndrome | 3/172 | 49/8085 | 0.0857 | 0.0857 | 0.411739879 | 950/23607/64423 | 3 |
| WP5300 | WP5300 | TROP2 regulatory signaling | 3/172 | 49/8085 | 0.0857 | 0.0857 | 0.411739879 | 5747/3678/8650 | 3 |
| WP2261 | WP2261 | Glioblastoma signaling pathways | 4/172 | 83/8085 | 0.1002 | 0.1002 | 0.472542808 | 5286/5583/5335/2033 | 4 |
| WP4726 | WP4726 | Sphingolipid metabolism: integrated pathway | 2/172 | 26/8085 | 0.1049 | 0.1049 | 0.486341206 | 8613/10558 | 2 |
| WP3932 | WP3932 | Focal adhesion: PI3K-Akt-mTOR-signaling pathway | 10/172 | 302/8085 | 0.1098 | 0.1098 | 0.500299271 | 1435/2034/3791/5747/5286/80310/3678/2321/10161/3643 | 10 |
| WP5036 | WP5036 | Angiotensin II receptor type 1 pathway | 2/172 | 28/8085 | 0.1188 | 0.1188 | 0.523043523 | 7048/80310 | 2 |
| WP5284 | WP5284 | Cell interactions of the pancreatic cancer microenvironment | 2/172 | 28/8085 | 0.1188 | 0.1188 | 0.523043523 | 1435/3791 | 2 |
| WP4239 | WP4239 | Epithelial to mesenchymal transition in colorectal cancer | 6/172 | 162/8085 | 0.1311 | 0.1311 | 0.549308366 | 5928/8503/7048/4040/3678/3728 | 6 |
| WP35 | WP35 | G protein signaling pathways | 4/172 | 92/8085 | 0.1322 | 0.1322 | 0.549308366 | 23683/11215/196883/5583 | 4 |
| WP1422 | WP1422 | Sphingolipid pathway | 2/172 | 30/8085 | 0.1331 | 0.1331 | 0.549308366 | 10558/10087 | 2 |
| WP2870 | WP2870 | Extracellular vesicle-mediated signaling in recipient cells | 2/172 | 30/8085 | 0.1331 | 0.1331 | 0.549308366 | 7048/324 | 2 |
| WP2355 | WP2355 | Corticotropin-releasing hormone signaling pathway | 4/172 | 93/8085 | 0.1360 | 0.1360 | 0.552630495 | 5747/7099/3728/5335 | 4 |

| | | | | | | | | | |
|---|---|---|---|---|---|---|---|---|---|
| WP3414 | WP3414 | Initiation of transcription and translation elongation at the HIV-1 LTR | 2/172 | 32/8085 | 0.1477 | 0.1477 | 0.553934466 | 51564/2033 | 2 |
| WP5072 | WP5072 | Modulators of TCR signaling and T cell activation | 3/172 | 63/8085 | 0.1499 | 0.1499 | 0.553934466 | 8525/8065/5335 | 3 |
| WP5222 | WP5222 | 2q13 copy number variation syndrome | 3/172 | 63/8085 | 0.1499 | 0.1499 | 0.553934466 | 1435/5747/2621 | 3 |
| WP474 | WP474 | Endochondral ossification | 3/172 | 64/8085 | 0.1550 | 0.1550 | 0.553934466 | 249/871/9510 | 3 |
| WP4808 | WP4808 | Endochondral ossification with skeletal dysplasias | 3/172 | 64/8085 | 0.1550 | 0.1550 | 0.553934466 | 249/871/9510 | 3 |
| WP4786 | WP4786 | Type I collagen synthesis in the context of osteogenesis imperfecta | 2/172 | 33/8085 | 0.1551 | 0.1551 | 0.553934466 | 871/4040 | 2 |
| WP4880 | WP4880 | Host-pathogen interaction of human coronaviruses - interferon induction | 2/172 | 33/8085 | 0.1551 | 0.1551 | 0.553934466 | 5610/10379 | 2 |
| WP4950 | WP4950 | 16p11.2 distal deletion syndrome | 2/172 | 33/8085 | 0.1551 | 0.1551 | 0.553934466 | 3791/3643 | 2 |
| WP5088 | WP5088 | Prostaglandin signaling | 2/172 | 33/8085 | 0.1551 | 0.1551 | 0.553934466 | 1435/196 | 2 |
| WP4874 | WP4874 | CAMKK2 pathway | 2/172 | 34/8085 | 0.1627 | 0.1627 | 0.565472208 | 1131/2033 | 2 |
| WP5117 | WP5117 | Cohesin complex - Cornelia de Lange syndrome | 2/172 | 34/8085 | 0.1627 | 0.1627 | 0.565472208 | 324/23047 | 2 |
| WP399 | WP399 | Wnt signaling pathway and pluripotency | 4/172 | 102/8085 | 0.1720 | 0.1720 | 0.573105498 | 324/4040/5583/2033 | 4 |
| WP4630 | WP4630 | Measles virus infection | 5/172 | 139/8085 | 0.1742 | 0.1742 | 0.573105498 | 8503/103/5610/7099/10379 | 5 |
| WP3303 | WP3303 | RAC1/PAK1/p38/MMP2 pathway | 3/172 | 68/8085 | 0.1758 | 0.1758 | 0.573105498 | 7075/5747/5829 | 3 |
| WP4685 | WP4685 | Melanoma | 3/172 | 68/8085 | 0.1758 | 0.1758 | 0.573105498 | 8503/3791/80243 | 3 |
| WP5083 | WP5083 | Neuroinflammation and glutamatergic signaling | 5/172 | 140/8085 | 0.1777 | 0.1777 | 0.573105498 | 84230/2744/7048/3597/3643 | 5 |
| WP3850 | WP3850 | Factors and pathways affecting insulin-like growth factor (IGF1)-Akt signaling | 2/172 | 36/8085 | 0.1779 | 0.1779 | 0.573105498 | 3488/253260 | 2 |
| WP2858 | WP2858 | Ectoderm differentiation | 5/172 | 142/8085 | 0.1849 | 0.1849 | 0.579208639 | 6498/4204/5787/3728/8675 | 5 |

| | | | | | | | | | |
|---|---|---|---|---|---|---|---|---|---|
| WP4331 | WP4331 | Neovascularisation processes | 2/172 | 37/8085 | 0.1856 | 0.1856 | 0.579208639 | 3791/4093 | 2 |
| WP4906 | WP4906 | 3q29 copy number variation syndrome | 3/172 | 71/8085 | 0.1919 | 0.1919 | 0.579208639 | 8503/9857/5829 | 3 |
| WP2380 | WP2380 | Brain-derived neurotrophic factor (BDNF) signaling pathway | 5/172 | 144/8085 | 0.1923 | 0.1923 | 0.579208639 | 249/57498/324/6197/5335 | 5 |
| WP4829 | WP4829 | mBDNF and proBDNF regulation of GABA neurotransmission | 2/172 | 38/8085 | 0.1933 | 0.1933 | 0.579208639 | 8503/5335 | 2 |
| WP524 | WP524 | G13 signaling pathway | 2/172 | 38/8085 | 0.1933 | 0.1933 | 0.579208639 | 10188/5585 | 2 |
| WP4673 | WP4673 | Male infertility | 5/172 | 145/8085 | 0.1960 | 0.1960 | 0.579208639 | 23369/196/9101/3643/2033 | 5 |
| WP2359 | WP2359 | Parkin-ubiquitin proteasomal system pathway | 3/172 | 72/8085 | 0.1973 | 0.1973 | 0.579208639 | 5707/25897/3309 | 3 |
| WP4674 | WP4674 | Head and neck squamous cell carcinoma | 3/172 | 74/8085 | 0.2083 | 0.2083 | 0.599038556 | 7048/253260/8650 | 3 |
| WP3937 | WP3937 | Microglia pathogen phagocytosis pathway | 2/172 | 40/8085 | 0.2089 | 0.2089 | 0.599038556 | 8503/5286 | 2 |
| WP710 | WP710 | DNA damage response (only ATM dependent) | 4/172 | 111/8085 | 0.2109 | 0.2109 | 0.599038556 | 8503/324/5286/3643 | 4 |
| WP2118 | WP2118 | Arrhythmogenic right ventricular cardiomyopathy | 3/172 | 75/8085 | 0.2138 | 0.2138 | 0.601026849 | 3672/3678/3728 | 3 |
| WP5055 | WP5055 | Burn wound healing | 4/172 | 113/8085 | 0.2198 | 0.2198 | 0.611370006 | 3791/7099/871/23476 | 4 |
| WP4565 | WP4565 | Neural crest cell migration in cancer | 2/172 | 43/8085 | 0.2325 | 0.2325 | 0.636917476 | 8503/57498 | 2 |
| WP4577 | WP4577 | Neurodegeneration with brain iron accumulation (NBIA) subtypes pathway | 2/172 | 44/8085 | 0.2404 | 0.2404 | 0.636917476 | 4204/10558 | 2 |
| WP4823 | WP4823 | Genes controlling nephrogenesis | 2/172 | 44/8085 | 0.2404 | 0.2404 | 0.636917476 | 3791/23607 | 2 |
| WP5205 | WP5205 | Clock-controlled autophagy in bone metabolism | 3/172 | 80/8085 | 0.2420 | 0.2420 | 0.636917476 | 1435/659/4093 | 3 |
| WP289 | WP289 | Myometrial relaxation and contraction pathways | 5/172 | 157/8085 | 0.2422 | 0.2422 | 0.636917476 | 3488/8525/196883/5583/5335 | 5 |
| WP268 | WP268 | Notch signaling | 2/172 | 45/8085 | 0.2483 | 0.2483 | 0.636917476 | 3955/8650 | 2 |

| | | | | | | | | | |
|---|---|---|---|---|---|---|---|---|---|
| WP2873 | WP2873 | Aryl hydrocarbon receptor pathway | 2/172 | 45/8085 | 0.2483 | 0.2483 | 0.636917476 | 196/2033 | 2 |
| WP5283 | WP5283 | Chronic hyperglycemia impairment of neuron function | 2/172 | 45/8085 | 0.2483 | 0.2483 | 0.636917476 | 23683/5583 | 2 |
| WP2038 | WP2038 | Microtubule cytoskeleton regulation | 2/172 | 46/8085 | 0.2562 | 0.2562 | 0.65093373 | 324/57551 | 2 |
| WP4971 | WP4971 | Phosphoinositides metabolism | 2/172 | 49/8085 | 0.2800 | 0.2800 | 0.703915716 | 5286/5335 | 2 |
| WP4559 | WP4559 | Interactions between immune cells and microRNAs in tumor microenvironment | 2/172 | 50/8085 | 0.2879 | 0.2879 | 0.703915716 | 7048/7099 | 2 |
| WP363 | WP363 | Wnt signaling pathway | 2/172 | 51/8085 | 0.2959 | 0.2959 | 0.703915716 | 324/4040 | 2 |
| WP4539 | WP4539 | Synaptic signaling pathways associated with autism spectrum disorder | 2/172 | 51/8085 | 0.2959 | 0.2959 | 0.703915716 | 8503/85358 | 2 |
| WP3286 | WP3286 | Copper homeostasis | 2/172 | 52/8085 | 0.3038 | 0.3038 | 0.703915716 | 324/54739 | 2 |
| WP673 | WP673 | ErbB signaling pathway | 3/172 | 92/8085 | 0.3115 | 0.3115 | 0.703915716 | 8503/5747/5335 | 3 |
| WP585 | WP585 | Interferon type I signaling pathways | 2/172 | 54/8085 | 0.3195 | 0.3195 | 0.703915716 | 2889/10379 | 2 |
| WP2018 | WP2018 | RANKL/RANK signaling pathway | 2/172 | 55/8085 | 0.3274 | 0.3274 | 0.703915716 | 5747/5335 | 2 |
| WP5124 | WP5124 | Alzheimer's disease | 7/172 | 264/8085 | 0.3306 | 0.3306 | 0.703915716 | 8503/1435/5610/5707/324/4040/3643 | 7 |
| WP4879 | WP4879 | Overlap between signal transduction pathways contributing to LMNA laminopathies | 2/172 | 57/8085 | 0.3430 | 0.3430 | 0.703915716 | 324/23405 | 2 |
| WP5265 | WP5265 | Neurogenesis regulation in the olfactory epithelium | 2/172 | 57/8085 | 0.3430 | 0.3430 | 0.703915716 | 4204/8650 | 2 |
| WP23 | WP23 | B cell receptor signaling pathway | 3/172 | 98/8085 | 0.3466 | 0.3466 | 0.703915716 | 2969/2889/5335 | 3 |
| WP2507 | WP2507 | Nanomaterial induced apoptosis | 1/172 | 20/8085 | 0.3499 | 0.3499 | 0.703915716 | 8837 | 1 |
| WP3640 | WP3640 | Imatinib and chronic myeloid leukemia | 1/172 | 20/8085 | 0.3499 | 0.3499 | 0.703915716 | 2321 | 1 |
| WP5223 | WP5223 | 2q21.1 copy number variation syndrome | 1/172 | 20/8085 | 0.3499 | 0.3499 | 0.703915716 | 324 | 1 |
| WP4223 | WP4223 | Ras signaling | 5/172 | 184/8085 | 0.3540 | 0.3540 | 0.703915716 | 8503/3791/2321/5335/3643 | 5 |

| | | | | | | | | | |
|---|---|---|---|---|---|---|---|---|---|
| WP5236 | WP5236 | Markers of kidney cell lineage | 2/172 | 59/8085 | 0.3585 | 0.3585 | 0.703915716 | 3791/79633 | 2 |
| WP3657 | WP3657 | Hematopoietic stem cell gene regulation by GABP alpha/beta complex | 1/172 | 21/8085 | 0.3637 | 0.3637 | 0.703915716 | 2033 | 1 |
| WP5053 | WP5053 | Development of ureteric collection system | 2/172 | 60/8085 | 0.3662 | 0.3662 | 0.703915716 | 659/79633 | 2 |
| WP558 | WP558 | Complement and coagulation cascades | 2/172 | 60/8085 | 0.3662 | 0.3662 | 0.703915716 | 2157/7056 | 2 |
| WP3945 | WP3945 | TYROBP causal network in microglia | 2/172 | 61/8085 | 0.3739 | 0.3739 | 0.703915716 | 375035/3597 | 2 |
| WP1584 | WP1584 | Type II diabetes mellitus | 1/172 | 22/8085 | 0.3773 | 0.3773 | 0.703915716 | 3643 | 1 |
| WP272 | WP272 | Blood clotting cascade | 1/172 | 22/8085 | 0.3773 | 0.3773 | 0.703915716 | 2157 | 1 |
| WP3599 | WP3599 | Transcription factor regulation in adipogenesis | 1/172 | 22/8085 | 0.3773 | 0.3773 | 0.703915716 | 3643 | 1 |
| WP4722 | WP4722 | Glycerolipids and glycerophospholipids | 1/172 | 22/8085 | 0.3773 | 0.3773 | 0.703915716 | 8525 | 1 |
| WP5111 | WP5111 | Familial hyperlipidemia type 4 | 1/172 | 22/8085 | 0.3773 | 0.3773 | 0.703915716 | 6400 | 1 |
| WP5194 | WP5194 | Synthesis of ceramides and 1-deoxyceramides | 1/172 | 22/8085 | 0.3773 | 0.3773 | 0.703915716 | 10558 | 1 |
| WP5180 | WP5180 | DYRK1A | 2/172 | 62/8085 | 0.3815 | 0.3815 | 0.703915716 | 5928/5978 | 2 |
| WP1544 | WP1544 | MicroRNAs in cardiomyocyte hypertrophy | 3/172 | 104/8085 | 0.3816 | 0.3816 | 0.703915716 | 8503/4040/51564 | 3 |
| WP4155 | WP4155 | Endometrial cancer | 2/172 | 63/8085 | 0.3891 | 0.3891 | 0.703915716 | 8503/324 | 2 |
| WP2509 | WP2509 | Nanoparticle triggered autophagic cell death | 1/172 | 23/8085 | 0.3906 | 0.3906 | 0.703915716 | 3643 | 1 |
| WP4197 | WP4197 | Immune response to tuberculosis | 1/172 | 23/8085 | 0.3906 | 0.3906 | 0.703915716 | 10379 | 1 |
| WP4357 | WP4357 | NRF2-ARE regulation | 1/172 | 23/8085 | 0.3906 | 0.3906 | 0.703915716 | 3643 | 1 |
| WP4790 | WP4790 | FGF23 signaling in hypophosphatemic rickets and related disorders | 1/172 | 23/8085 | 0.3906 | 0.3906 | 0.703915716 | 249 | 1 |
| WP4830 | WP4830 | GDNF/RET signaling axis | 1/172 | 23/8085 | 0.3906 | 0.3906 | 0.703915716 | 79633 | 1 |
| WP4942 | WP4942 | 15q13.3 copy number variation syndrome | 1/172 | 23/8085 | 0.3906 | 0.3906 | 0.703915716 | 871 | 1 |

| | | | | | | | | | |
|---|---|---|---|---|---|---|---|---|---|
| WP3624 | WP3624 | Lung fibrosis | 2/172 | 64/8085 | 0.3966 | 0.3966 | 0.703915716 | 6498/4204 | 2 |
| WP1528 | WP1528 | Physiological and pathological hypertrophy of the heart | 1/172 | 24/8085 | 0.4036 | 0.4036 | 0.703915716 | 3977 | 1 |
| WP325 | WP325 | Triacylglyceride synthesis | 1/172 | 24/8085 | 0.4036 | 0.4036 | 0.703915716 | 8613 | 1 |
| WP334 | WP334 | GPCRs, class B secretin-like | 1/172 | 24/8085 | 0.4036 | 0.4036 | 0.703915716 | 23266 | 1 |
| WP5272 | WP5272 | LDL- influence on CD14 and TLR4 | 1/172 | 24/8085 | 0.4036 | 0.4036 | 0.703915716 | 7099 | 1 |
| WP4666 | WP4666 | Hepatitis B infection | 4/172 | 152/8085 | 0.4060 | 0.4060 | 0.703915716 | 8503/7048/7099/2033 | 4 |
| WP536 | WP536 | Calcium regulation in cardiac cells | 4/172 | 152/8085 | 0.4060 | 0.4060 | 0.703915716 | 1131/196883/5583/10052 | 4 |
| WP2324 | WP2324 | AGE/RAGE pathway | 2/172 | 66/8085 | 0.4116 | 0.4116 | 0.703915716 | 249/3643 | 2 |
| WP2857 | WP2857 | Mesodermal commitment pathway | 4/172 | 154/8085 | 0.4156 | 0.4156 | 0.703915716 | 23499/659/6926/4091 | 4 |
| WP2895 | WP2895 | Differentiation of white and brown adipocyte | 1/172 | 25/8085 | 0.4163 | 0.4163 | 0.703915716 | 4093 | 1 |
| WP3668 | WP3668 | Hypothesized pathways in pathogenesis of cardiovascular disease | 1/172 | 25/8085 | 0.4163 | 0.4163 | 0.703915716 | 7048 | 1 |
| WP4585 | WP4585 | Cancer immunotherapy by PD-1 blockade | 1/172 | 25/8085 | 0.4163 | 0.4163 | 0.703915716 | 10725 | 1 |
| WP4842 | WP4842 | Mammalian disorder of sexual development | 1/172 | 25/8085 | 0.4163 | 0.4163 | 0.703915716 | 23543 | 1 |
| WP1403 | WP1403 | AMP-activated protein kinase signaling | 2/172 | 67/8085 | 0.4190 | 0.4190 | 0.703915716 | 8503/3643 | 2 |
| WP5293 | WP5293 | Acute myeloid leukemia | 2/172 | 67/8085 | 0.4190 | 0.4190 | 0.703915716 | 8503/3728 | 2 |
| WP3613 | WP3613 | Photodynamic therapy-induced unfolded protein response | 1/172 | 27/8085 | 0.4410 | 0.4410 | 0.703915716 | 3309 | 1 |
| WP3845 | WP3845 | Canonical and non-canonical Notch signaling | 1/172 | 27/8085 | 0.4410 | 0.4410 | 0.703915716 | 84441 | 1 |
| WP176 | WP176 | Folate metabolism | 2/172 | 70/8085 | 0.4410 | 0.4410 | 0.703915716 | 1435/3643 | 2 |
| WP5130 | WP5130 | Th17 cell differentiation pathway | 2/172 | 70/8085 | 0.4410 | 0.4410 | 0.703915716 | 196/5335 | 2 |

| | | | | | | | | | |
|---|---|---|---|---|---|---|---|---|---|
| WP2880 | WP2880 | Glucocorticoid receptor pathway | 2/172 | 71/8085 | 0.4482 | 0.4482 | 0.703915716 | 6397/7056 | 2 |
| WP3851 | WP3851 | TLR4 signaling and tolerance | 1/172 | 28/8085 | 0.4529 | 0.4529 | 0.703915716 | 7099 | 1 |
| WP5121 | WP5121 | Sphingolipid metabolism in senescence | 1/172 | 28/8085 | 0.4529 | 0.4529 | 0.703915716 | 10558 | 1 |
| WP5200 | WP5200 | Dravet syndrome | 1/172 | 28/8085 | 0.4529 | 0.4529 | 0.703915716 | 253260 | 1 |
| WP4255 | WP4255 | Non-small cell lung cancer | 2/172 | 72/8085 | 0.4554 | 0.4554 | 0.703915716 | 8503/5335 | 2 |
| WP4216 | WP4216 | Chromosomal and microsatellite instability in colorectal cancer | 2/172 | 73/8085 | 0.4625 | 0.4625 | 0.703915716 | 7048/324 | 2 |
| WP2361 | WP2361 | Gastric cancer network 1 | 1/172 | 29/8085 | 0.4646 | 0.4646 | 0.703915716 | 324 | 1 |
| WP3869 | WP3869 | Cannabinoid receptor signaling | 1/172 | 29/8085 | 0.4646 | 0.4646 | 0.703915716 | 196 | 1 |
| WP179 | WP179 | Cell cycle | 3/172 | 120/8085 | 0.4722 | 0.4722 | 0.703915716 | 9184/51433/2033 | 3 |
| WP3929 | WP3929 | Chemokine signaling pathway | 4/172 | 166/8085 | 0.4725 | 0.4725 | 0.703915716 | 8503/5747/5829/196883 | 4 |
| WP2583 | WP2583 | T cell receptor and co-stimulatory signaling | 1/172 | 30/8085 | 0.4760 | 0.4760 | 0.703915716 | 5335 | 1 |
| WP4941 | WP4941 | GPR143 in melanocytes and retinal pigment epithelium cells | 1/172 | 30/8085 | 0.4760 | 0.4760 | 0.703915716 | 196883 | 1 |
| WP530 | WP530 | Cytokines and inflammatory response | 1/172 | 30/8085 | 0.4760 | 0.4760 | 0.703915716 | 1435 | 1 |
| WP4341 | WP4341 | Non-genomic actions of 1,25 dihydroxyvitamin D3 | 2/172 | 75/8085 | 0.4766 | 0.4766 | 0.703915716 | 7099/5583 | 2 |
| WP4549 | WP4549 | Fragile X syndrome | 3/172 | 122/8085 | 0.4831 | 0.4831 | 0.703915716 | 4204/23405/5335 | 3 |
| WP2034 | WP2034 | Leptin signaling pathway | 2/172 | 76/8085 | 0.4835 | 0.4835 | 0.703915716 | 5747/5335 | 2 |
| WP2037 | WP2037 | Prolactin signaling pathway | 2/172 | 76/8085 | 0.4835 | 0.4835 | 0.703915716 | 5747/5829 | 2 |
| WP5218 | WP5218 | Extrafollicular and follicular B cell activation by SARS-CoV-2 | 2/172 | 76/8085 | 0.4835 | 0.4835 | 0.703915716 | 7099/283131 | 2 |
| WP3858 | WP3858 | Toll-like receptor signaling related to MyD88 | 1/172 | 31/8085 | 0.4872 | 0.4872 | 0.703915716 | 7099 | 1 |
| WP3996 | WP3996 | Ethanol effects on histone modifications | 1/172 | 31/8085 | 0.4872 | 0.4872 | 0.703915716 | 51564 | 1 |

| | | | | | | | | | |
|---|---|---|---|---|---|---|---|---|---|
| WP4204 | WP4204 | Tumor suppressor activity of SMARCB1 | 1/172 | 31/8085 | 0.4872 | 0.4872 | 0.703915716 | 5928 | 1 |
| WP4912 | WP4912 | SARS coronavirus and innate immunity | 1/172 | 31/8085 | 0.4872 | 0.4872 | 0.703915716 | 10379 | 1 |
| WP5122 | WP5122 | Prostaglandin and leukotriene metabolism in senescence | 1/172 | 31/8085 | 0.4872 | 0.4872 | 0.703915716 | 3488 | 1 |
| WP4949 | WP4949 | 16p11.2 proximal deletion syndrome | 2/172 | 77/8085 | 0.4904 | 0.4904 | 0.703915716 | 58508/2186 | 2 |
| WP4538 | WP4538 | Regulatory circuits of the STAT3 signaling pathway | 2/172 | 78/8085 | 0.4973 | 0.4973 | 0.703915716 | 3977/253260 | 2 |
| WP2363 | WP2363 | Gastric cancer network 2 | 1/172 | 32/8085 | 0.4981 | 0.4981 | 0.703915716 | 157769 | 1 |
| WP3893 | WP3893 | Development and heterogeneity of the ILC family | 1/172 | 32/8085 | 0.4981 | 0.4981 | 0.703915716 | 196 | 1 |
| WP244 | WP244 | Alpha 6 beta 4 signaling pathway | 1/172 | 33/8085 | 0.5089 | 0.5089 | 0.703915716 | 5747 | 1 |
| WP3676 | WP3676 | BDNF-TrkB signaling | 1/172 | 33/8085 | 0.5089 | 0.5089 | 0.703915716 | 5335 | 1 |
| WP4481 | WP4481 | Resistin as a regulator of inflammation | 1/172 | 33/8085 | 0.5089 | 0.5089 | 0.703915716 | 5335 | 1 |
| WP465 | WP465 | Tryptophan metabolism | 1/172 | 33/8085 | 0.5089 | 0.5089 | 0.703915716 | 196 | 1 |
| WP4900 | WP4900 | Purinergic signaling | 1/172 | 33/8085 | 0.5089 | 0.5089 | 0.703915716 | 10161 | 1 |
| WP3971 | WP3971 | OSX and miRNAs in tooth development | 1/172 | 34/8085 | 0.5194 | 0.5194 | 0.710983321 | 249 | 1 |
| WP58 | WP58 | Monoamine GPCRs | 1/172 | 34/8085 | 0.5194 | 0.5194 | 0.710983321 | 1131 | 1 |
| WP3617 | WP3617 | Photodynamic therapy-induced NF-kB survival signaling | 1/172 | 35/8085 | 0.5296 | 0.5296 | 0.718748139 | 8837 | 1 |
| WP1471 | WP1471 | Target of rapamycin signaling | 1/172 | 36/8085 | 0.5397 | 0.5397 | 0.718748139 | 253260 | 1 |
| WP4877 | WP4877 | Host-pathogen interaction of human coronaviruses - MAPK signaling | 1/172 | 36/8085 | 0.5397 | 0.5397 | 0.718748139 | 6197 | 1 |
| WP34 | WP34 | Ovarian infertility | 1/172 | 37/8085 | 0.5495 | 0.5495 | 0.718748139 | 23224 | 1 |
| WP4150 | WP4150 | Wnt signaling in kidney disease | 1/172 | 37/8085 | 0.5495 | 0.5495 | 0.718748139 | 4040 | 1 |
| WP4320 | WP4320 | Effect of progerin on genes involved in Hutchinson-Gilford progeria syndrome | 1/172 | 37/8085 | 0.5495 | 0.5495 | 0.718748139 | 5928 | 1 |

| | | | | | | | | | |
|---|---|---|---|---|---|---|---|---|---|
| WP4558 | WP4558 | Overview of interferons-mediated signaling pathway | 1/172 | 37/8085 | 0.5495 | 0.5495 | 0.718748139 | 10379 | 1 |
| WP4698 | WP4698 | Vitamin D-sensitive calcium signaling in depression | 1/172 | 37/8085 | 0.5495 | 0.5495 | 0.718748139 | 493 | 1 |
| WP5269 | WP5269 | Genetic causes of porto-sinusoidal vascular disease | 1/172 | 37/8085 | 0.5495 | 0.5495 | 0.718748139 | 2969 | 1 |
| WP2059 | WP2059 | Alzheimer's disease and miRNA effects | 7/172 | 329/8085 | 0.5548 | 0.5548 | 0.720229412 | 8503/1435/5610/5707/324/4040/3643 | 7 |
| WP15 | WP15 | Selenium micronutrient network | 2/172 | 87/8085 | 0.5561 | 0.5561 | 0.720229412 | 57190/3643 | 2 |
| WP5098 | WP5098 | T-cell activation SARS-CoV-2 | 2/172 | 88/8085 | 0.5623 | 0.5623 | 0.722449005 | 253260/5335 | 2 |
| WP4263 | WP4263 | Pancreatic adenocarcinoma pathway | 2/172 | 89/8085 | 0.5685 | 0.5685 | 0.722449005 | 8503/7048 | 2 |
| WP4871 | WP4871 | Kisspeptin/kisspeptin receptor system in the ovary | 1/172 | 39/8085 | 0.5686 | 0.5686 | 0.722449005 | 5583 | 1 |
| WP4336 | WP4336 | ncRNAs involved in Wnt signaling in hepatocellular carcinoma | 2/172 | 90/8085 | 0.5746 | 0.5746 | 0.722449005 | 324/4040 | 2 |
| WP2526 | WP2526 | PDGF pathway | 1/172 | 40/8085 | 0.5778 | 0.5778 | 0.722449005 | 5335 | 1 |
| WP3941 | WP3941 | Oxidative damage response | 1/172 | 40/8085 | 0.5778 | 0.5778 | 0.722449005 | 10188 | 1 |
| WP4564 | WP4564 | Neural crest cell migration during development | 1/172 | 40/8085 | 0.5778 | 0.5778 | 0.722449005 | 8503 | 1 |
| WP138 | WP138 | Androgen receptor signaling pathway | 2/172 | 91/8085 | 0.5806 | 0.5806 | 0.722449005 | 5747/2033 | 2 |
| WP2877 | WP2877 | Vitamin D receptor pathway | 4/172 | 191/8085 | 0.5834 | 0.5834 | 0.722449005 | 8853/3488/7056/634 | 4 |
| WP1433 | WP1433 | Nucleotide-binding oligomerization domain (NOD) pathway | 1/172 | 41/8085 | 0.5868 | 0.5868 | 0.722449005 | 55914 | 1 |
| WP4290 | WP4290 | Metabolic reprogramming in colon cancer | 1/172 | 42/8085 | 0.5956 | 0.5956 | 0.722449005 | 2744 | 1 |
| WP2853 | WP2853 | Endoderm differentiation | 3/172 | 145/8085 | 0.6002 | 0.6002 | 0.722449005 | 324/1112/2186 | 3 |
| WP5224 | WP5224 | 2q37 copy number variation syndrome | 3/172 | 145/8085 | 0.6002 | 0.6002 | 0.722449005 | 8837/3597/4735 | 3 |

| | | | | | | | | | |
|---|---|---|---|---|---|---|---|---|---|
| WP2911 | WP2911 | miRNA targets in ECM and membrane receptors | 1/172 | 43/8085 | 0.6043 | 0.6043 | 0.722449005 | 3672 | 1 |
| WP4136 | WP4136 | Fibrin complement receptor 3 signaling pathway | 1/172 | 43/8085 | 0.6043 | 0.6043 | 0.722449005 | 7099 | 1 |
| WP4249 | WP4249 | Hedgehog signaling pathway | 1/172 | 43/8085 | 0.6043 | 0.6043 | 0.722449005 | 64750 | 1 |
| WP5198 | WP5198 | Inflammatory bowel disease signaling | 1/172 | 43/8085 | 0.6043 | 0.6043 | 0.722449005 | 7099 | 1 |
| WP4658 | WP4658 | Small cell lung | 2/172 | 96/8085 | 0.6099 | 0.6099 | 0.725893637 | 8503/5747 | 2 |
| WP314 | WP314 | Fas ligand pathway and stress induction of heat shock proteins | 1/172 | 44/8085 | 0.6128 | 0.6128 | 0.725996864 | 8837 | 1 |
| WP3878 | WP3878 | ATM signaling in development and disease | 1/172 | 45/8085 | 0.6210 | 0.6210 | 0.731969328 | 55183 | 1 |
| WP4258 | WP4258 | lncRNA in canonical Wnt signaling and colorectal cancer | 2/172 | 99/8085 | 0.6267 | 0.6267 | 0.731969328 | 324/4040 | 2 |
| WP4566 | WP4566 | Translation inhibitors in chronically activated PDGFRA cells | 1/172 | 46/8085 | 0.6291 | 0.6291 | 0.731969328 | 8503 | 1 |
| WP5078 | WP5078 | T cell modulation in pancreatic cancer | 1/172 | 46/8085 | 0.6291 | 0.6291 | 0.731969328 | 953 | 1 |
| WP1591 | WP1591 | Heart development | 1/172 | 47/8085 | 0.6371 | 0.6371 | 0.731969328 | 659 | 1 |
| WP1541 | WP1541 | Energy metabolism | 1/172 | 48/8085 | 0.6448 | 0.6448 | 0.731969328 | 2033 | 1 |
| WP2848 | WP2848 | Pluripotent stem cell differentiation pathway | 1/172 | 48/8085 | 0.6448 | 0.6448 | 0.731969328 | 1435 | 1 |
| WP410 | WP410 | Exercise-induced circadian regulation | 1/172 | 48/8085 | 0.6448 | 0.6448 | 0.731969328 | 5813 | 1 |
| WP4262 | WP4262 | Breast cancer pathway | 3/172 | 155/8085 | 0.6454 | 0.6454 | 0.731969328 | 8503/324/4040 | 3 |
| WP117 | WP117 | GPCRs, other | 2/172 | 103/8085 | 0.6483 | 0.6483 | 0.731969328 | 23266/1131 | 2 |
| WP75 | WP75 | Toll-like receptor signaling pathway | 2/172 | 103/8085 | 0.6483 | 0.6483 | 0.731969328 | 8503/7099 | 2 |
| WP107 | WP107 | Translation factors | 1/172 | 50/8085 | 0.6599 | 0.6599 | 0.738769431 | 5610 | 1 |
| WP286 | WP286 | IL-3 signaling pathway | 1/172 | 50/8085 | 0.6599 | 0.6599 | 0.738769431 | 2889 | 1 |
| WP706 | WP706 | Sudden infant death syndrome (SIDS) susceptibility pathways | 3/172 | 160/8085 | 0.6666 | 0.6666 | 0.739235817 | 5978/4204/2033 | 3 |
| WP4932 | WP4932 | 7q11.23 copy number variation syndrome | 2/172 | 107/8085 | 0.6688 | 0.6688 | 0.739235817 | 2969/23476 | 2 |

| | | | | | | | | | |
|---|---|---|---|---|---|---|---|---|---|
| WP5090 | WP5090 | Complement system in neuronal development and plasticity | 2/172 | 107/8085 | 0.6688 | 0.6688 | 0.739235817 | 8547/2621 | 2 |
| WP4321 | WP4321 | Thermogenesis | 2/172 | 108/8085 | 0.6738 | 0.6738 | 0.739235817 | 6197/196883 | 2 |
| WP1533 | WP1533 | Vitamin B12 metabolism | 1/172 | 52/8085 | 0.6743 | 0.6743 | 0.739235817 | 3643 | 1 |
| WP4352 | WP4352 | Ciliary landscape | 4/172 | 216/8085 | 0.6798 | 0.6798 | 0.740734186 | 324/2664/23011/10640 | 4 |
| WP2406 | WP2406 | Cardiac progenitor differentiation | 1/172 | 53/8085 | 0.6813 | 0.6813 | 0.740734186 | 3791 | 1 |
| WP2795 | WP2795 | Cardiac hypertrophic response | 1/172 | 54/8085 | 0.6881 | 0.6881 | 0.742047977 | 51564 | 1 |
| WP395 | WP395 | IL-4 signaling pathway | 1/172 | 54/8085 | 0.6881 | 0.6881 | 0.742047977 | 2033 | 1 |
| WP195 | WP195 | IL-1 signaling pathway | 1/172 | 55/8085 | 0.6948 | 0.6948 | 0.743185036 | 5335 | 1 |
| WP3959 | WP3959 | DNA IR-double strand breaks and cellular response via ATM | 1/172 | 55/8085 | 0.6948 | 0.6948 | 0.743185036 | 55183 | 1 |
| WP4222 | WP4222 | Phosphodiesterases in neuronal function | 1/172 | 56/8085 | 0.7013 | 0.7013 | 0.744556951 | 196883 | 1 |
| WP4754 | WP4754 | IL-18 signaling pathway | 5/172 | 275/8085 | 0.7017 | 0.7017 | 0.744556951 | 8837/1112/7052/5335/5585 | 5 |
| WP428 | WP428 | Wnt signaling | 2/172 | 115/8085 | 0.7070 | 0.7070 | 0.7472046 | 324/4040 | 2 |
| WP5115 | WP5115 | Network map of SARS-CoV-2 signaling pathway | 4/172 | 226/8085 | 0.7138 | 0.7138 | 0.748568291 | 7048/196/51028/10379 | 4 |
| WP2272 | WP2272 | Pathogenic Escherichia coli infection | 1/172 | 58/8085 | 0.7140 | 0.7140 | 0.748568291 | 7099 | 1 |
| WP3527 | WP3527 | Pre-implantation embryo | 1/172 | 59/8085 | 0.7201 | 0.7201 | 0.75200999 | 6926 | 1 |
| WP3865 | WP3865 | Novel intracellular components of RIG-I-like receptor pathway | 1/172 | 60/8085 | 0.7261 | 0.7261 | 0.755288759 | 10521 | 1 |
| WP3863 | WP3863 | T-cell antigen receptor (TCR) pathway during Staphylococcus aureus infection | 1/172 | 62/8085 | 0.7377 | 0.7377 | 0.764362295 | 5335 | 1 |
| WP183 | WP183 | Proteasome degradation | 1/172 | 64/8085 | 0.7488 | 0.7488 | 0.769852118 | 5707 | 1 |
| WP5097 | WP5097 | CCL18 signaling pathway | 1/172 | 64/8085 | 0.7488 | 0.7488 | 0.769852118 | 2033 | 1 |
| WP4329 | WP4329 | miRNA role in immune response in sepsis | 1/172 | 65/8085 | 0.7542 | 0.7542 | 0.772383595 | 7099 | 1 |

| | | | | | | | | | |
|---|---|---|---|---|---|---|---|---|---|
| WP4657 | WP4657 | 22q11.2 copy number variation syndrome | 2/172 | 129/8085 | 0.7650 | 0.7650 | 0.780419514 | 3842/3837 | 2 |
| WP5039 | WP5039 | SARS-CoV-2 innate immunity evasion and cell-specific immune response | 1/172 | 68/8085 | 0.7697 | 0.7697 | 0.782170788 | 2033 | 1 |
| WP382 | WP382 | MAPK signaling pathway | 4/172 | 248/8085 | 0.7791 | 0.7791 | 0.788642334 | 7048/9693/6197/57551 | 4 |
| WP1982 | WP1982 | Sterol regulatory element-binding proteins (SREBP) signaling | 1/172 | 71/8085 | 0.7842 | 0.7842 | 0.790830937 | 3837 | 1 |
| WP4655 | WP4655 | Cytosolic DNA-sensing pathway | 1/172 | 74/8085 | 0.7978 | 0.7978 | 0.801486024 | 103 | 1 |
| WP4656 | WP4656 | Joubert syndrome | 1/172 | 76/8085 | 0.8064 | 0.8064 | 0.804064797 | 4646 | 1 |
| WP2882 | WP2882 | Nuclear receptors meta-pathway | 5/172 | 315/8085 | 0.8065 | 0.8065 | 0.804064797 | 7048/196/6397/7056/2033 | 5 |
| WP4022 | WP4022 | Pyrimidine metabolism | 1/172 | 84/8085 | 0.8373 | 0.8373 | 0.823235484 | 953 | 1 |
| WP4396 | WP4396 | Nonalcoholic fatty liver disease | 2/172 | 155/8085 | 0.8468 | 0.8468 | 0.823235484 | 8503/3643 | 2 |
| WP2840 | WP2840 | Hair follicle development: cytodifferentiation - part 3 of 3 | 1/172 | 87/8085 | 0.8476 | 0.8476 | 0.823235484 | 3488 | 1 |
| WP4298 | WP4298 | Acute viral myocarditis | 1/172 | 87/8085 | 0.8476 | 0.8476 | 0.823235484 | 7099 | 1 |
| WP254 | WP254 | Apoptosis | 1/172 | 88/8085 | 0.8508 | 0.8508 | 0.823235484 | 8837 | 1 |
| WP477 | WP477 | Cytoplasmic ribosomal proteins | 1/172 | 88/8085 | 0.8508 | 0.8508 | 0.823235484 | 6197 | 1 |
| WP5234 | WP5234 | Bardet-Biedl syndrome | 1/172 | 88/8085 | 0.8508 | 0.8508 | 0.823235484 | 8239 | 1 |
| WP2446 | WP2446 | Retinoblastoma gene in cancer | 1/172 | 90/8085 | 0.8572 | 0.8572 | 0.823235484 | 5928 | 1 |
| WP1772 | WP1772 | Apoptosis modulation and signaling | 1/172 | 92/8085 | 0.8633 | 0.8633 | 0.823235484 | 8837 | 1 |
| WP3925 | WP3925 | Amino acid metabolism | 1/172 | 92/8085 | 0.8633 | 0.8633 | 0.823235484 | 2744 | 1 |
| WP69 | WP69 | T-cell receptor signaling pathway | 1/172 | 92/8085 | 0.8633 | 0.8633 | 0.823235484 | 5335 | 1 |
| WP231 | WP231 | TNF-alpha signaling pathway | 1/172 | 93/8085 | 0.8662 | 0.8662 | 0.823235484 | 8837 | 1 |
| WP4746 | WP4746 | Thyroid hormones production and peripheral downstream signaling effects | 1/172 | 93/8085 | 0.8662 | 0.8662 | 0.823235484 | 9728 | 1 |

| | | | | | | | | | |
|---|---|---|---|---|---|---|---|---|---|
| WP4963 | WP4963 | p53 transcriptional gene network | 1/172 | 96/8085 | 0.8747 | 0.8747 | 0.828291739 | 10379 | 1 |
| WP2064 | WP2064 | Neural crest differentiation | 1/172 | 101/8085 | 0.8876 | 0.8876 | 0.837532038 | 51564 | 1 |
| WP4536 | WP4536 | Genes related to primary cilium development (based on CRISPR) | 1/172 | 104/8085 | 0.8947 | 0.8947 | 0.841237486 | 57534 | 1 |
| WP2431 | WP2431 | Spinal cord injury | 1/172 | 121/8085 | 0.9273 | 0.9273 | 0.868818458 | 7099 | 1 |
| WP2806 | WP2806 | Complement system | 1/172 | 124/8085 | 0.9319 | 0.9319 | 0.87005129 | 22918 | 1 |
| WP5095 | WP5095 | Overview of proinflammatory and profibrotic mediators | 1/172 | 129/8085 | 0.9390 | 0.9390 | 0.873541437 | 1435 | 1 |
| WP2884 | WP2884 | NRF2 pathway | 1/172 | 141/8085 | 0.9531 | 0.9531 | 0.883536842 | 7048 | 1 |
| WP455 | WP455 | GPCRs, class A rhodopsin-like | 2/172 | 263/8085 | 0.9780 | 0.9780 | 0.901003644 | 1131/10161 | 2 |
| WP4803 | WP4803 | Ciliopathies | 1/172 | 184/8085 | 0.9817 | 0.9817 | 0.901003644 | 8239 | 1 |
| WP702 | WP702 | Metapathway biotransformation Phase I and II | 1/172 | 185/8085 | 0.9821 | 0.9821 | 0.901003644 | 9394 | 1 |
